# Likely community transmission of COVID-19 infections between neighboring, persistent hotspots in Ontario, Canada

**DOI:** 10.1101/2021.12.06.21267360

**Authors:** Eliseos J. Mucaki, Ben C. Shirley, Peter K. Rogan

**Author notes:** Correspondence: Peter K. Rogan, Ph.D., Departments of Biochemistry and Oncology, Schulich School of Medicine and Dentistry, University of Western Ontario, London ON N6A 2C1 Canada.

## Abstract

**Introduction:** This study aimed to produce community-level geo-spatial mapping of confirmed COVID-19 cases in Ontario, Canada in near real-time to support decision-making. This was accomplished by area-to-area geostatistical analysis, space-time integration, and spatial interpolation of COVID-19 positive individuals.

**Methods:** COVID-19 cases and locations were curated for geostatistical analyses from March 2020 through June 2021, corresponding to the first, second, and third waves of infections. Daily cases were aggregated according to designated forward sortation area [FSA], and postal codes [PC] in municipal regions covering Hamilton, Kitchener/Waterloo, London, Ottawa, Toronto, and Windsor/Essex county. Hotspots were identified with area-to-area tests including Getis-Ord Gi^*^, Global Moran’s I spatial autocorrelation, and Local Moran’s I asymmetric clustering and outlier analyses. Case counts were also interpolated across geographic regions by Empirical Bayesian Kriging, which localizes high concentrations of COVID-19 positive tests, independent of FSA or PC boundaries. The *Geostatistical Disease Epidemiology Toolbox*, which is freely-available software, automates the identification of these regions and produces digital maps for public health professionals to assist in pandemic management of contact tracing and distribution of other resources.

**Results/Discussion:** This study provided indicators in real-time of likely, community-level disease transmission through innovative geospatial analyses of COVID-19 incidence data. Municipal and provincial results were validated by comparisons with known outbreaks at long-term care and other high density residences and on farms. PC-level analyses revealed hotspots at higher geospatial resolution than public reports of FSAs, and often sooner. Results of different tests and kriging were compared to determine consistency among hotspot assignments. Concurrent or consecutive hotspots in close proximity suggested potential community transmission of COVID-19 from cluster and outlier analysis of neighboring PCs and by kriging. Results were also stratified by population based-categories (sex, age, and presence/absence of comorbidities). Earlier recognition of hotspots could reduce public health burdens of COVID-19 and expedite contact tracing.

## Introduction

Many critical elements of disease epidemiology were unknown during the early stages of the COVID-19 pandemic. This included the case fatality rate, at what point during the natural history of the disease was it most transmissible (before, during or after the onset of symptoms), the frequencies of asymptomatic presentation of positive cases, and the duration of infectivity^1^. Quarantining, social distancing, and isolation of infected populations were quickly recognized as effective public health measures. Without implementation of these measures in Ontario, COVID-19 patients presenting with severe disease would have exceeded available hospital capacity^2^. Delayed testing and the sheer numbers of infected individuals often precluded comprehensive contact tracing^3^. Indeed, COVID-19 testing was limited in many regions within Canada early in the pandemic, as was the availability of necessary resources (e.g., ICUs, nursing and other clinical personnel, personal protective equipment)^4,5^. Limited resources must be adequately allocated to those locations where the rates of transmission of COVID-19 are high and where the highest numbers of infected individuals reside. This highlights the need for an effective, systematic way of tracking concentrations of this disease after community-level transmission. We address the persistence of geographic disease hotspots as a potential sentinel for efficient distribution of sparse resources.

The SARS-CoV-2 virus spreads by person-to-person contact, especially through respiration^6^. Geographic epidemiology can be a key factor for the prevention of disease spread, as it can be used to monitor disease progression (or regression) and manage allocation of medical resources. Geostatistical methods have been utilized to map communicable diseases such as influenza-like diseases^7^, dengue fever^8^, cholera^9^, legionellosis and shigellosis, among others^10^. Various geostatistical analyses have been used in the surveillance of COVID-19 spread across the globe^11–17^, often with the geo-spatial software, ArcGIS (ESRI)^18^. Other available tools for geostatistical epidemiology include SaTScan^TM 19^, GeoDa^20^, and R statistical software environment^21^, however these were not used in the present study because of their requirement to input disease background levels, which was not relevant for COVID-19. Available case data analyzed has been typically aggregated at the municipal^11,16^ or county level^13,14,22^, however higher resolution spatial analyses has been limited as finer granularity for the distribution of COVID-19 cases has often been lacking^16,18^. We performed geostatistical area-to-area analyses as well as interpolation of COVID-19 cases within six municipalities of the Canadian province of Ontario (Hamilton, Kitchener/Waterloo, London, Ottawa, Toronto and Windsor/Essex region) distributed at the level of individual forward sortation areas (FSA) and postal codes (PCs). Area-to-area analyses were performed on COVID-19 case data at the provincial level by FSA, a geographical unit which can encompass hundreds of PCs. Each PC comprises an average of 19 households (Statistics Canada, 2006).

The success of short-term COVID-19 geostatistical studies at the county level early in the pandemic^22^ suggested that hotspots and localized concentrations of infected individuals could be delineated more precisely with epidemiological data collected with higher granularity. The present analysis evaluates the three major waves of COVID-19 infection to date in Ontario, allowing for the observation of trends and commonalities between waves. We utilized ArcGIS to carry out multiple area-to-area geostatistical tests, including Getis-Ord Gi* hotspot analysis (Figure 1)^23,24^, Cluster and Outlier analysis (Figures 2A and 2B; Anselin Local Moran’s I^25^), and Spatial Autocorrelation (Figures 2C and 2D; Global Moran’s I^26^). Hotspots were also inferred from spatially-interpolated counts by kriging and represented as topographic contour maps^27,28^ utilizing the ArcGIS geographic information system. Area-to-Area Gi* analysis finds regions with a high disease burden (i.e., model for community-driven spread), but does not inform how these cases are distributed throughout the region. PC-level point-to-point analyses (e.g., kriging) can be employed to provide context of how cases cluster and identify localized hotspots, where cases are concentrated in a single PC and independent of the disease burden of the FSA it is within (Figure 3). The Getis Ord Gi* local statistic identifies regions with high disease burden with high, spatially clustered COVID-19 case counts (a local sum) relative to the rest of the province^23^. Space-time Gi* analysis also uses this statistic to find hotspots that are both spatially and temporally clustered with other neighboring features (Figure 1D). Cluster and Outlier analysis identifies high-case regions that are geospatially clustered with other neighboring regions with a similar rate of cases (high-case clusters; Figure 2A), or those with neighboring regions with significantly lower COVID-19 case counts (high-case outliers; Figure 2B). The Global Moran’s I index determines if the distribution of cases is clustered, dispersed, or random within a region^26^. The Moran’s I index is positive for spatially clustered cases between postal codes, which is compared to the expected value (i.e., if the spatial relationship was random) to compute a z-score and significance level. Kriging is a geostatistical interpolation method which predicts values over a non-sampled region by analyzing a subset of locations with known values^28^. As a combination of geostatistical methods were applied to daily COVID-19 case data at the provincial [FSAs] and municipality [PCs] level for >450 days, efficient scripts were necessary to automate these analyses.

**Figure 1.**
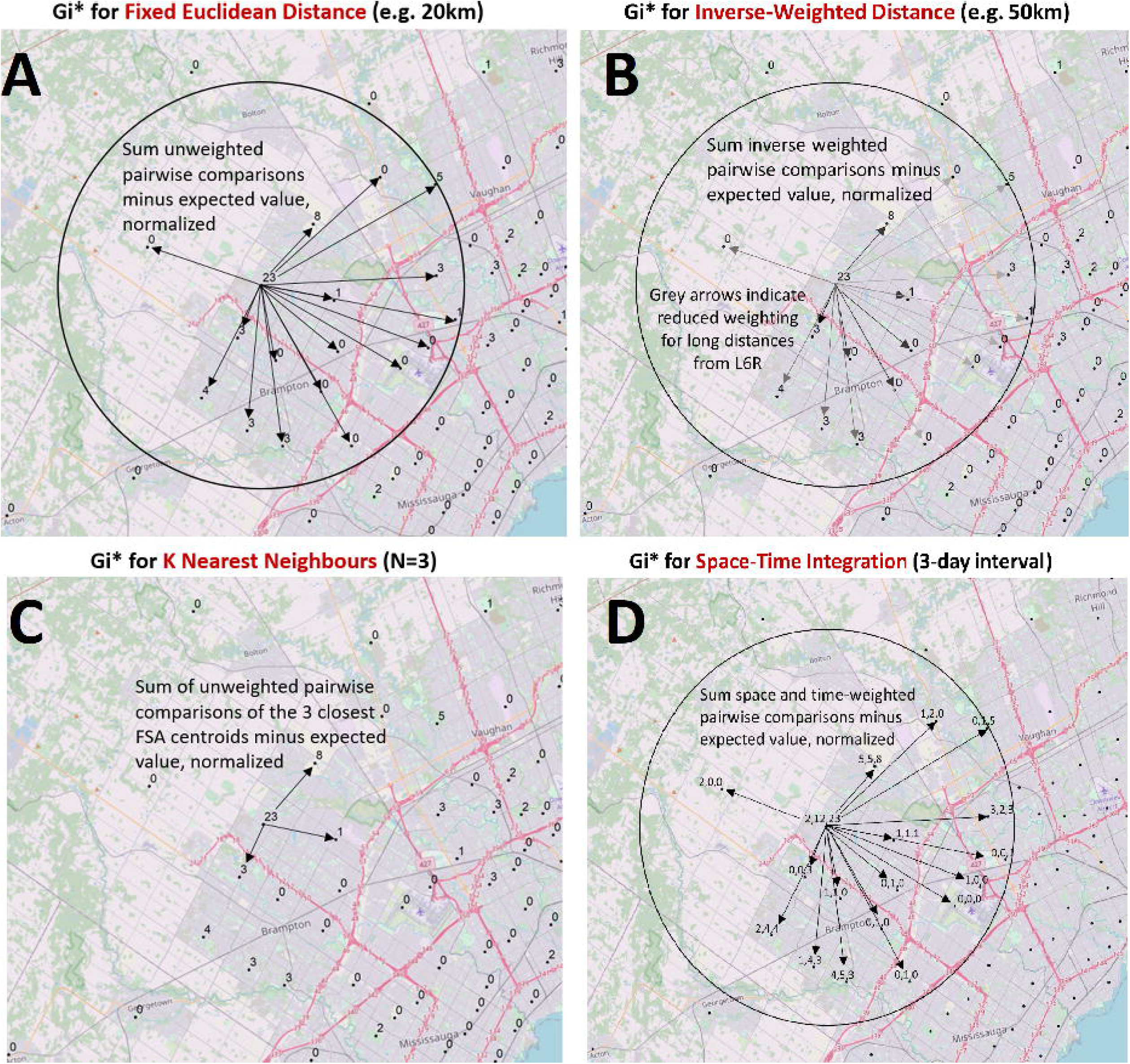
Modeling Spatial Relationships by Area-to-Area Geostatistical Analyses. Case counts aggregated at the centroids of FSAs or PCs over daily-or multi-day windows are compared to cases in neighboring FSAs or PCs and evaluated with geostatistical tests. These spatial relationships are defined by modeling spatial interactions between the FSA or PC being evaluated and the neighbors being tested. The panels describing these interactions indicate: (A) The fixed distance band weighs all neighbors within a specified distance equally, (B) The inverse distance band is based on distance decay where neighbors further from the target feature is weighted less relative to closer neighbors, (C) K-Nearest Neighbors constructs a spatial relationship which assesses a feature with a specified number of its most proximate neighbors based on centroid distances (here, N=3), (D) Space-Time integration defines feature relationships in terms of both a space (fixed distance) and a time window. Features that are proximate to one another in space and time are analyzed together, as relationships are assessed relative to the location and time stamp of the target feature. Area-to-area geostatistical tests used include Gi* analysis, Cluster and Outlier analysis, and Spatial Autocorrelation.

**Figure 2.**
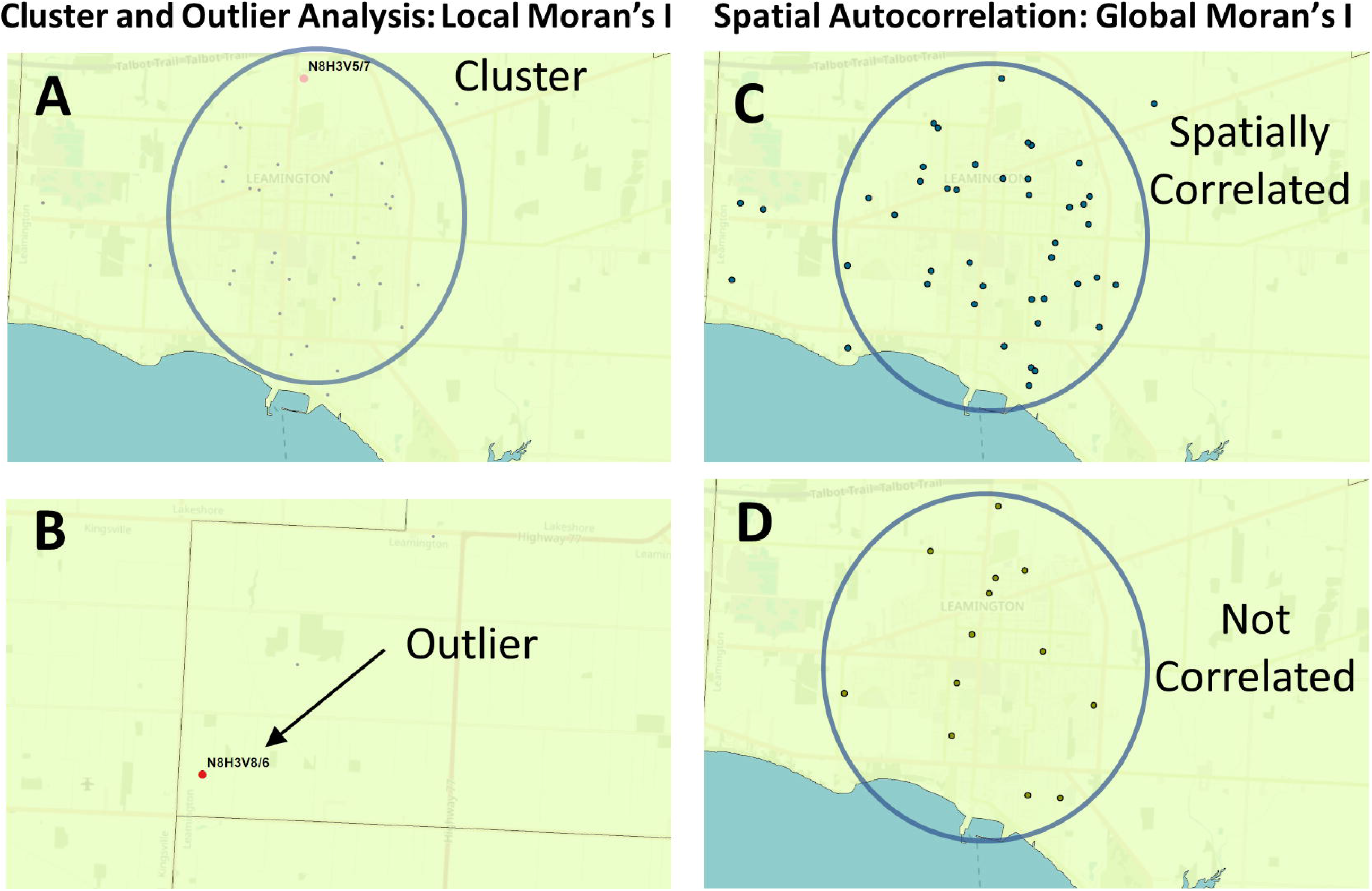
Geostatistical Analysis of the Distribution of COVID-19 Cases. The Global and Local Moran’s I statistics determine whether the numbers of cases in different PCs or FSAs in a region are distinct from one another or are related. Local Moran’s I (also known as Cluster and Outlier analysis) computes a Moran’s I value, or spatial autocorrelation, between counts within sets of FSAs or PCs. Global Moran’s I evaluates the overall case clustering in a region. Panels shown indicate these relationships in Leamington FSA N8H: A) A positive local Moran’s I index indicates that the regions neighboring N8H 3V5 has similar attributes (a high-case cluster; pink dot). B) A negative local Moran’s I index demonstrates that cases the region surrounding the target are dissimilar (a high-case outlier in N8H 3V8; red dot). C) The Global Moran’s I (Spatial Autocorrelation) index demonstrates whether cases are clustered, dispersed, or randomly distributed. In this example, cases were significantly clustered from Dec. 21 to 23, 2020 (p=0.03; p-value signifies whether the null hypothesis [a random distribution] can be rejected). D) The case distribution was not significantly clustered on Dec. 2 to 4, 2020 (p=0.7).

**Figure 3.**
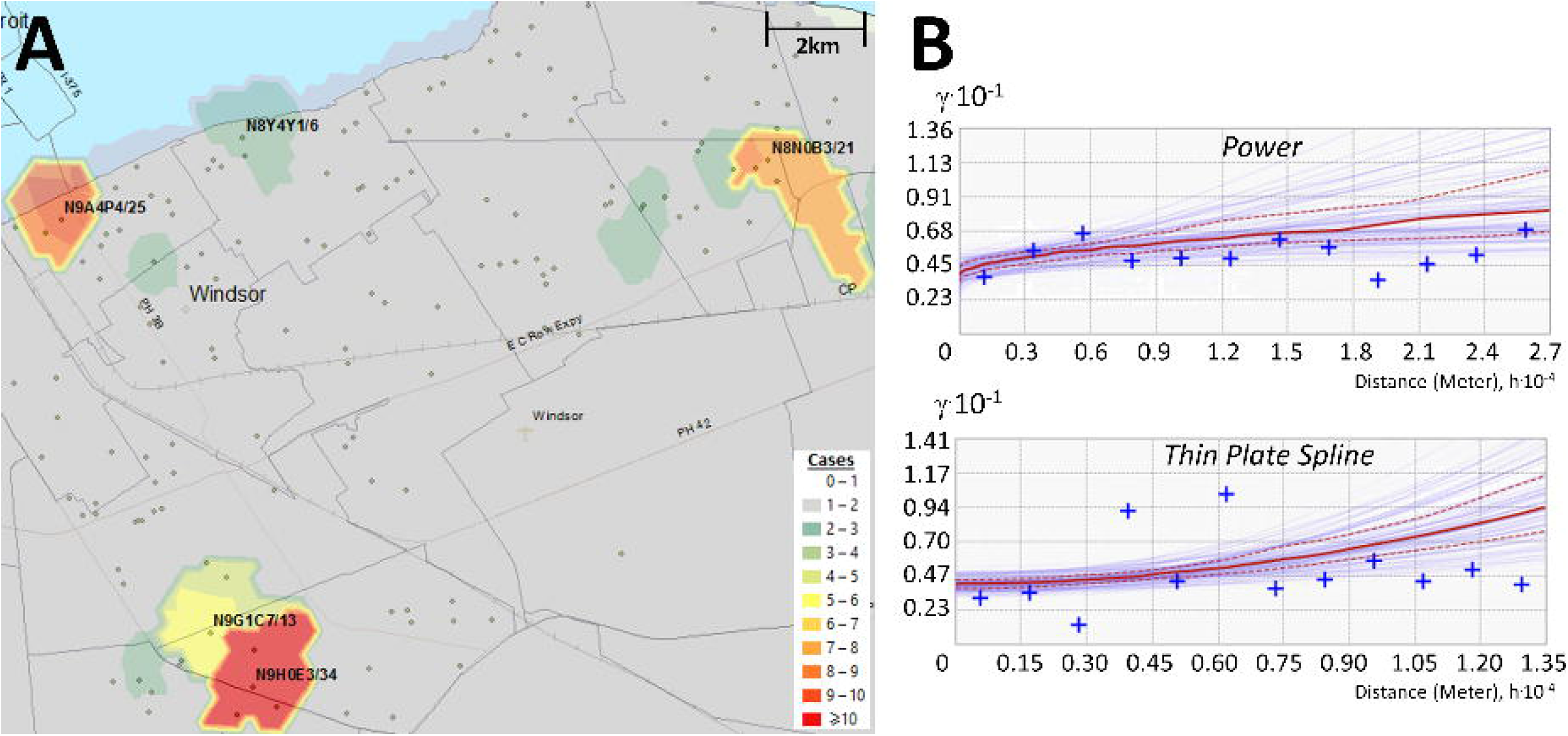
Interpolating the Distribution of COVID-19 Cases by Empirical Bayesian Kriging. Kriging estimates case counts by interpolating unsampled regions between counts at known locations. Panels indicate: (A) Topographic contours were generated by Empirical Bayesian kriging (EBK) when analyzing PC-level case data in Windsor (Dec. 18 to 20, 2020) and interpolate different case levels within the region; (B) EBK uses empirical semivariograms which describe the correlations between the distance separating a pair of locations with COVID-19-positive individuals, ‘h’, and their corresponding semivariance ‘*γ*’. Larger values of *γ* indicate lower spatial autocorrelation between numbers of cases. A semivariogram model fits a curve through *γ* vs. distance that predicts *γ* at unsampled locations between known data points. The ‘Power’ semivariogram (B, top) is a parabolic mathematical model based on distance, while the Thin Plate Spline semivariogram (B, bottom) is a non-rigid model that passes exactly through bins of paired locations (grouped by distance and direction; blue crosses) while minimizing surface curvature. Red lines represent the median of the distribution of semivariogram models.

These geostatistical tools were used to identify and krige COVID-19 hotspots, clusters, and outliers within Ontario at a high level of granularity. With the results of these analyses, we can ask questions regarding the distribution of hotspots relative to others in both time and space. For example, this study evaluates significant levels of COVID-19 over a series of consecutive days which we term ‘streaks’. This allowed us to determine the true size and persistence of hotspots, compare and contrast geostatistical analyses of COVID-19 waves highlighting recurrent hotspots and clusters of PCs with high case counts. We also investigated the order in the progression of infection across a region in a statistically robust way. Empirical Bayesian Kriging (EBK) identified concentrations of COVID-19 that transcended the boundaries of FSAs and PCs. Ultimately, consistency between the results obtained by these different approaches reinforces the conclusions that might be drawn from them based on implementation as independent geostatistical tests.

## Methods

### Geostatistical Analysis

COVID-19 cases from March 26^th^, 2020 to June 25^th^, 2021 at the FSA (Ontario-wide) and PC-level (for 6 populated areas: Hamilton, Kitchener/Waterloo, London Ottawa, Toronto, and Windsor, including Essex county) were evaluated using ArcGIS Desktop 10.7 [ESRI] with the *Geostatistical Analyst* extension. Daily cases compiled by FSA were evaluated with Cluster and Outlier analysis, Gi* ‘hotspot’ analysis, and with space-time integrated Gi* analysis. Cluster and Outlier analysis computes a Local Moran’s I index score for each feature. A positive score indicates that a feature is clustered with other similar high-case count regions, while a negative index indicates that a feature is an outlier. The z-score and p-value indicates the statistical significance of the index value. A false discovery rate (FDR) correction was applied to this and Gi* analysis, which changes the threshold of significance of p-value ≤ 0.05 to a reduced value within the 95% confidence interval [C.I.], thereby accounting for multiple testing and spatial dependency. Z-scores are also determined in Gi* analysis, with higher positive scores indicating greater clustering of cases in a region. Space-time Gi* analysis was based on a single day interval but incorporated temporal trends based on the days preceding and subsequent to each date. We set a minimum threshold of 10 confirmed cases for an FSA to be considered significant by these three approaches, as this threshold was found to provide reliable and conservative detection of known hotspots using the Gi* test. Over two thirds of FSAs with ≥ 10 cases between March 1^st^ and Sept. 24^th^, 2020 were found to be significant by Gi* [99% C.I.] using both a fixed and inverse distance metric (Extended data^29^, Section 1 -Figure S1). Spatial relationships were evaluated to determine which distance metric best reflected the relationship between the test location and its surrounding FSAs by fixed distance (all FSAs within a given threshold distance [10km] are equally weighted with the test FSA), inverse distance (FSAs closer to the test FSA are weighted more highly), and by K Nearest Neighbors (KNN; comparing counts in the test FSA and its closest K neighbors).

PC-level case data (binned over 3-day sliding windows) for each municipality were evaluated by Spatial Autocorrelation (Global Moran’s I), Cluster and Outlier analysis, and EBK. Spatial Autocorrelation used FDR correction to account for multiple testing; it was not applied for PC-level Cluster and Outlier analysis due a limitation of the method in high-case count regions where all locations were deemed non-significant. The KNN distance metric was used to define the spatial relationship between features (with K=3) for all PCs, including those with zero cases. PCs with ≥6 cases were reported if deemed significant by Cluster and Outlier analysis. This is consistent with case minimums used in other COVID-19 geostatistical studies^13,14^. Kriging generates a topographic contour map of the number of COVID-19 cases interpolated over the evaluated region, as indicated below.

COVID-19 testing data compiled by ICES contained metadata on tested individuals, such as gender, age, and relevant predicate health conditions. To identify geospatially significant cases in these groups, case data was stratified by gender (Male: 241,393; Female: 247,152; those without gender information were ignored), by age (≥ 65 years old: 63,746; < 65 years: 424,854), and by presence or absence of at least one chronic health condition (187,242; includes chronic asthma, congestive heart failure, chronic obstructive pulmonary disease [COPD], dementia, hypertension and/or diabetes). 301,358 patients were without one of these conditions. Individuals with these conditions are grouped as an “at-risk population”, consistent with provincial terminology. These case subsets were then evaluated with Gi* (FSA-level), Spatial Autocorrelation, Cluster and Outlier analysis and kriging (PC-level).

Results from PC-level Cluster and Outlier analysis were further analyzed with a program script to identify significantly, highly clustered PCs over consecutive periods (i.e., termed high-case cluster “streaks”; *Local_Morans_Analysis*.*Recurrent_Clustered_PC_Identifier*.*pl;* see Data Availability section^30^). We also identified which of these PC streaks occurred in close proximity (*Local_Morans_Analysis*.*Clustered_Streak_Pairing_Program*.*pl*) based on the neighbors of each postal code from PC centroid data (neighbors identified by *Ontario_City_Closest_Postal_Code_Identification*.*pl; see: FSA and PC boundary files* subsection). Directional network graphs which visualize the interconnectivity of these paired high-case cluster streaks were created in MATLAB using built-in *digraph* and *plot* functions, where each edge (the connection between two PCs) represents a single pair of PCs with nearly concomitant or overlapping streaks.

Software automation was required to computationally evaluate daily and windowed case data over 16 months at the geographic resolution of FSAs and PCs across multiple cities with different geostatistical tests. Using the ArcPy package (ESRI), which enables Python programs to access ArcGIS’s geoprocessing tools and extensions, program scripts were developed to perform geostatistical analyses across Ontario. These scripts set conditional variables which define the spatial relationship used (fixed, inverse or KNN), the threshold distance (or for KNN, the number of neighbors of the target feature), the distance method (how distances were calculated; set to ‘Euclidean’), row standardization (set to ‘none’), and whether FDR correction was used (applied to FSA-level Gi* and Cluster and Outlier analysis, and PC-level Spatial Autocorrelation analysis). ArcPy scripts read in case data (in tabular format), iterated sequentially through each date skipping those without positive cases, applied the associated geostatistical tool, and converted the output into text files. Tabular output was subsequently filtered by removing PCs without cases to avoid producing outputs of prohibitive size.

Additional ArcPy scripts imported and visualized the results from Gi*, kriging and Cluster and Outlier analysis onto maps displayed in the ArcGIS system. These scripts label regions above the prerequisite case minimums to identify the location (FSA or PC) and case count, symbolize the results, i.e., to assign a colour to a location which represents its significance based on the analysis performed, and convert the map to a PNG formatted image. To image results of kriging analyses, program scripts simultaneously visualized the locations of COVID-19 cases and the kriging layer by setting transparency to 30%. The extent of the region visualized was defined manually in ArcGIS prior to generation of map images.

These processes were then integrated and streamlined in a software package named the *Geostatistical Epidemiology Toolbox* (see Data Availability section^30^), which is accessed through the ArcMap graphical user interface. This resource provides the same geostatistical operations and map imaging developed for this project in a simplified, user-friendly environment. This toolbox bundles manual pre-processing, analysis, and post-analytical steps into a set of linked, uninterrupted workflows. For example, tabular case data is converted into shapefiles and, optionally, the ArcGIS window is aligned to the city of interest when performing map imaging. The user-defined parameters for these operations are set through an interface entirely within ArcMap, which then allows a series of tasks to be carried out automatically. No prior Python programming knowledge is required. While the toolbox was designed to evaluate COVID-19 case data in Ontario, the software can be easily modified to accommodate other Canadian provinces and, with additional effort, other countries. To enable kriging functionality within the toolbox, the *Advanced Geostatistical Analyst* module is required to be installed in ArcMap as well as access to PC and FSA boundary shapefiles. These shapefiles are also required for PC-level Local Moran’s I and Global Moran’s I analyses. This package is not bundled with any of the program scripts developed to evaluate geostatistical output (beyond data imaging), however these are provided in the accompanying Zenodo archive^30^. The toolbox is distributed under the GNU General Public License v3.0.

### Preparation of Ontario COVID-19 Case Data from ICES

Access to anonymized Ontario COVID-19 test results through the ICES Research and Analytic Environment (RAE) was obtained through Applied Health Research Questions (AHRQ) project #P0950.096 approved by the Ontario Ministry of Health and Long-Term Care (MOHLTC). ICES collects health care data for analysis and evaluation of the health system. Ontario-wide COVID-19 data tables maintained in ICES were pre-processed to ensure that only the chronologically first laboratory-confirmed viral RNA polymerase chain reaction (PCR) SARS-CoV-2 test of a specific individual was counted (while repeat infections occur, these are assumed to be significantly less frequent). PCs associated with each positive case were cross-referenced to their respective FSA (derived from the 2016 Canadian Census boundary file from Statistics Canada) to identify and filter out invalid entries. The PCs provided were sourced from the OLIS (Ontario Laboratories Information System) and the RPDB (Registered Persons) databases. The OLIS postal code was more likely to relate to a current address and was therefore prioritized. However, it is also the most error prone. Reasons for rejection include blank entries, invalid FSAs (e.g., out of province FSA), or intentional masking. The RPDB PC was used if the OLIS PC was rejected. Entries where both PCs were invalid were rare (<0.1% of all unique positive cases). Non-residents or those ineligible for the Ontario Health Insurance Plan (OHIP) are also not included. Cases were separately aggregated by FSAs and PCs, then converted into tables structured as rows (FSA or PC count) by columns (date) as input to ArcGIS Desktop. Cases were based on the date of sampling, rather than when test results were reported, to more accurately reflect the inception of infections.

As of June 2021, three separate and distinct waves of COVID-19 cases have been recognized in Ontario (and elsewhere). We delineated the inception and termination dates of each wave from the rates of change in infection rates (cases/day^2^) using the second derivative of daily case counts. Stochasticity in day-to-day variation due to testing and case reporting caused fluctuations in changes of daily counts, even when averaging cases over multiple consecutive days. Consecutive dates without fluctuations in counts (between -10 and +10 cases/day^2^) were rare. Therefore, dates were selected where the second derivative remained between -10 and +10 cases/day^2^ immediately preceding rapid increases in cases that defined inception of a wave, or after infections ceased decreasing in counts, defining termination of a wave. The first wave (1) was found to span from Mar. 26^th^ to Jun. 16^th^, 2020, second wave (2) began on Sept. 25^th^, 2020 and continued until Feb. 14^th^, 2021, and the third wave (3) covered Mar. 17^th^ to Jun. 23^rd^, 2021.

Of the geostatistical methods tested, PC-level counts of infected individuals were the most geospatially resolved granular COVID-19 case data. Many PCs comprise of small geographic areas with low population densities. This made them more prone to the effects of day-to-day stochasticity in patient ascertainment and laboratory reporting, especially when case counts were low. To address this, PC-level case data was also aggregated over multi-day sliding windows, effectively smoothing out this source of variation. Although 3 and 7-day windows were initially analyzed, longer windows were not as useful for determining the inception of hotspots, a goal of this analysis, and were eventually discounted. 3-day windows have been used in other geostatistical studies of COVID-19^13,14^.

Access to, pre-processing and geostatistical analysis of COVID-19 daily cases and associated metadata required the RAE computer system at ICES. To ensure patient privacy, ICES stipulated that the counts in all FSAs and/or PCs exhibiting between 1 and 5 daily COVID-19 cases be masked. We modified counts of FSAs meeting these criteria and assessed their impact on the geostatistical analysis. Gi* analysis of ground truth case data was compared with the analysis of two modified FSA-level case datasets (where all instances of between 1 and 5 case counts were either converted to a single count or randomized between 1 and 5). More than one third of the locations that were deemed significant hotspots by Gi* from actual case counts (from March 1^st^ to Sept. 24^th^, 2020) were not flagged when locations were masked and assigned a count of 1; greater than one half were missed when case counts were randomized (Extended data^29^, Section 1 - Table S1). Discordant hotspots identified from actual vs. masked case counts were reduced in FSAs with high-case counts (≥10) but were nevertheless unacceptably frequent (14% or 28% discordant, respectively, by masking low case counts as 1 or by randomization). We concluded that errors introduced by masking count values precluded analysis of masked data and required all work to be carried out within the RAE computing environment.

### FSA and PC boundary files

Shapefiles which store geospatial data [points, lines or polygons] with related attribute information and depict the 2016 census geographic boundaries of FSAs across Canada were obtained from Statistics Canada (https://www12.statcan.gc.ca/census-recensement/2011/geo/bound-limit/bound-limit-2016-eng.cfm). PC boundary shapefiles created by DMTI (“DMTI_2019_CMPCS_LocalDeliveryUnitsRegion.shp”) were accessed through the Scholar’s Geoportal at the University of Western Ontario (http://geo2.scholarsportal.info/). These files were used to compute latitude/longitude coordinates of the centroid for each FSA and PC (with the ArcGIS “Calculate Geometry” function), to validate PCs provided for each individual in the COVID-19 test dataset, and to convert spatial interpolation maps (such as those generated by kriging) into machine-readable text files (using ArcGIS “Intersection” function between the spatial map and with each boundary file). The ArcGIS “Split by Attribute” function was used to limit PC boundary files to the province of Ontario. This was necessary, both because data from other provinces were not available, and because of system limitations affecting processing and memory requirements for large geographic regions.

Minor discrepancies between the FSA and PC boundary files were identified which had little or no impact on our geostatistical analysis. Nearly 9% of all PCs in Ontario overlapped multiple FSAs. The majority of these PCs were present in multiple locations and occurred in rural areas. Approximately 2.7% of PCs intersected with an incorrect FSA, which was likely the result of asynchronous creation of the corresponding shapefiles. These locations rarely contained many COVID-19 cases (only 11 PCs had >5 cases over any 3-day span in 2020), including: N9H 0E3 (Windsor, ON) which overlaps with the FSA N9G; N8N 0B3 (Tecumseh, ON) which overlaps the bordering FSA N8R, and M3H 5V9 (Toronto, ON) which intersected with neighboring FSA M3J. These discrepancies might impact the interpolation of cases in FSAs based on kriging analysis. Since the discrepant PCs were typically found on the border of FSAs, these conflicts had minimal effect on kriging accuracy.

### Empirical Bayesian Kriging Analysis

Kriging estimates case counts in unsampled regions by interpolation between counts at known locations. These values are spatially correlated with degree of separation from centroids of PCs or FSA with known cases. A topographic contour map is generated which illustrates the interpolated case levels within the analyzed region. The spatial distribution of the disease outbreak was determined for Hamilton, Kitchener/Waterloo, London, Ottawa, Toronto, and Windsor/Essex county by kriging analysis of PC-level COVID-19 case data over consecutive, overlapping 3-day windows. Kriging was also performed for other municipalities bordering Toronto (Ajax, Brampton, Markham, Mississauga, Pickering, Richmond Hill and Vaughan; see results in Extended data^29^, Section 3). Only PCs with ≥1 case were included in this analysis, as locations without confirmed cases were found to severely depress kriging signals. Several types of kriging (Ordinary, Universal, Simple and EBK) were evaluated for accurate detection of high case clusters in municipalities with high and moderate population densities (Toronto and London, respectively).

Contour maps generated by EBK best matched regions of known cases used for analysis. Ordinary and Universal kriging yielded similar results for locations with high case counts (≥50) but lacked the sensitivity of EBK in regions with moderate cases. Simple kriging failed to define most contours correctly, regardless of case count. Therefore, EBK was chosen for all kriging analyses presented in this study.

Semivariograms describe how the distances separating COVID-19 positive individuals and the semivariance of counts at these locations are correlated. The best model that fits this relationship using empirically defined distance data was used to predict COVID-19 count levels at unsampled locations between the centroids of FSAs or PCs. Semivariogram models assessed included Power, Linear, Thin Plate Spline [TPS], Whittle, Exponential, K-Bessel. Kriging parameters for these models were considered, such as sector type, search radius, neighborhood type and the number of neighbors considered for each location. Through empirical testing, the ‘Power’ semivariogram model (based on a parabolic relationship to distance between locations) was chosen based on interpolated values that best represented actual case counts in low/moderately dense populations (Extended data^29^, Section 1 - Figure S2A). Accuracy was maximized when interpolated values were based on 3 neighboring locations with known counts. However, the performance of the ‘Power’ semivariogram to interpolate high-case regions in Toronto, which exhibited the highest case counts of any municipality, failed to model some localized hotspots. The TPS semivariogram model, a non-rigid representation that passes exactly through points while minimizing surface curvature, performed best in this densely populated region (Extended data^29^, Section 1 - Figure S2B). Power-based kriging contours were generally larger, while TPS-based kriging contours were smaller, bifurcated and more frequent. TPS, in some instances, generated kriging artifacts that did not correlate with known hotspots. Increasing the numbers of neighbors to 5 reduced the occurrence of these artifacts without negatively impacting the accuracy of the contour map. Consequently, we chose to evaluate cities with low-to-moderate population densities (Hamilton, Kitchener/Waterloo, London, Ottawa and Windsor/Essex) with the ‘Power’ semivariogram model (3 neighbors), whereas the Toronto metropolitan area was evaluated with the TPS semivariogram model (5 neighbors). All other EBK parameters were defaulted to those provided by the ArcGIS Advanced Geostatistics toolbox.

We have previously demonstrated that circumscribing the boundaries of analyzed regions by specifying these locations with zero counts improves kriging accuracy, especially at locations proximate to boundaries^31^. This approach was used in the present study of interpolated COVID-19 cases for each municipality in Ontario. Municipal borders were derived by combining all PCs associated with the region of interest. The optimal densities and distances of the zero-count buffer circumscribing municipalities by programmatically generating boundaries. Distances between the boundary to the city border were varied from 0.1, 0.5, 2, and 5km, and densities between locations of 0.05, 0.25, 0.5 and 1km. Kriging analysis was iterated for each of these defined zero-count borders, and the kriging layers generated were compared to the original kriging results. The location of each kriging contour properly coincided with regions of high case counts, regardless of the shape and density of the zero-count boundary locations. Raising the boundary density consistently increased the COVID-19 case value interpolated of a high-case region while simultaneously reducing the total area of the kriging contours around the region, regardless of semivariogram model used. This effect was less prominent in regions with higher density of cases (i.e., Toronto), and in regions further from the city boundaries (as the kriging contours exhibited consistent shape and magnitude >6 km away from the city boundary). Considering these observations, we chose to use a moderate value for each parameter (0.5km density and 1 km distance from the city boundary). The kriging tool in the *Geostatistical Epidemiology Toolbox*, however, automatically generates these boundaries 1 km from the selected border with 250m of distance between each location.

Kriging analysis generates a series of shapefiles containing contour maps that define the interpolated level of COVID-19 cases of a region. An ArcPy script intersected map contours derived from kriging and either FSA or PC boundary files, which associated each kriging-derived case count with these locations. The results of these intersections were displayed as distinct contour distributions (or kriging ‘symbologies’): one for instances with a wider range of inferred case count levels (0-1, 1-5, 5-10, 10-15 … 35-40, 40-50, >50 cases), and another for low densities of inferred cases, which was discriminated by single counts (0-1, 1-2 … 9-10, >10 cases). In compliance with ICES’s reporting criteria, we defined any interpolated value > 5 cases (either ≥5-6 or ≥5-10 contour, depending on the symbology type used) as significant, and display PC or FSA designations for only these locations. The intersected area (m^2^) between contours and the corresponding FSA or PC was determined to compute the size of localized hotspots (e.g., the total area of the FSA or PC considered significant by kriging).

## Results

Changes in the distribution of COVID-19 infections over time were evaluated through geostatistical analysis within both FSAs and PCs. PC-level analysis can map COVID-19 spread at a finer resolution, which more precisely identifies hotspots concentrated within individual postal codes (e.g., a localized hotspot). Area-to-area geostatistical analysis of cases in FSAs is better suited when COVID-19 cases are distributed across the entire FSA (e.g., an area with high disease burden by community-driven spread; Figure 4), rather than when cases are concentrated in a single location. This method does not, however, provide any information regarding the overall distribution of cases within the FSA. Spatial Autocorrelation analysis (Global Moran’s I) was therefore performed to identify FSAs where COVID-19 cases are clustered at the PC-level (potential evidence of disease transmission). These geostatistical methods combine analyses of COVID-19 cases at multiple levels of resolution, facilitating interpretation of the case distribution throughout the province.

**Figure 4.**
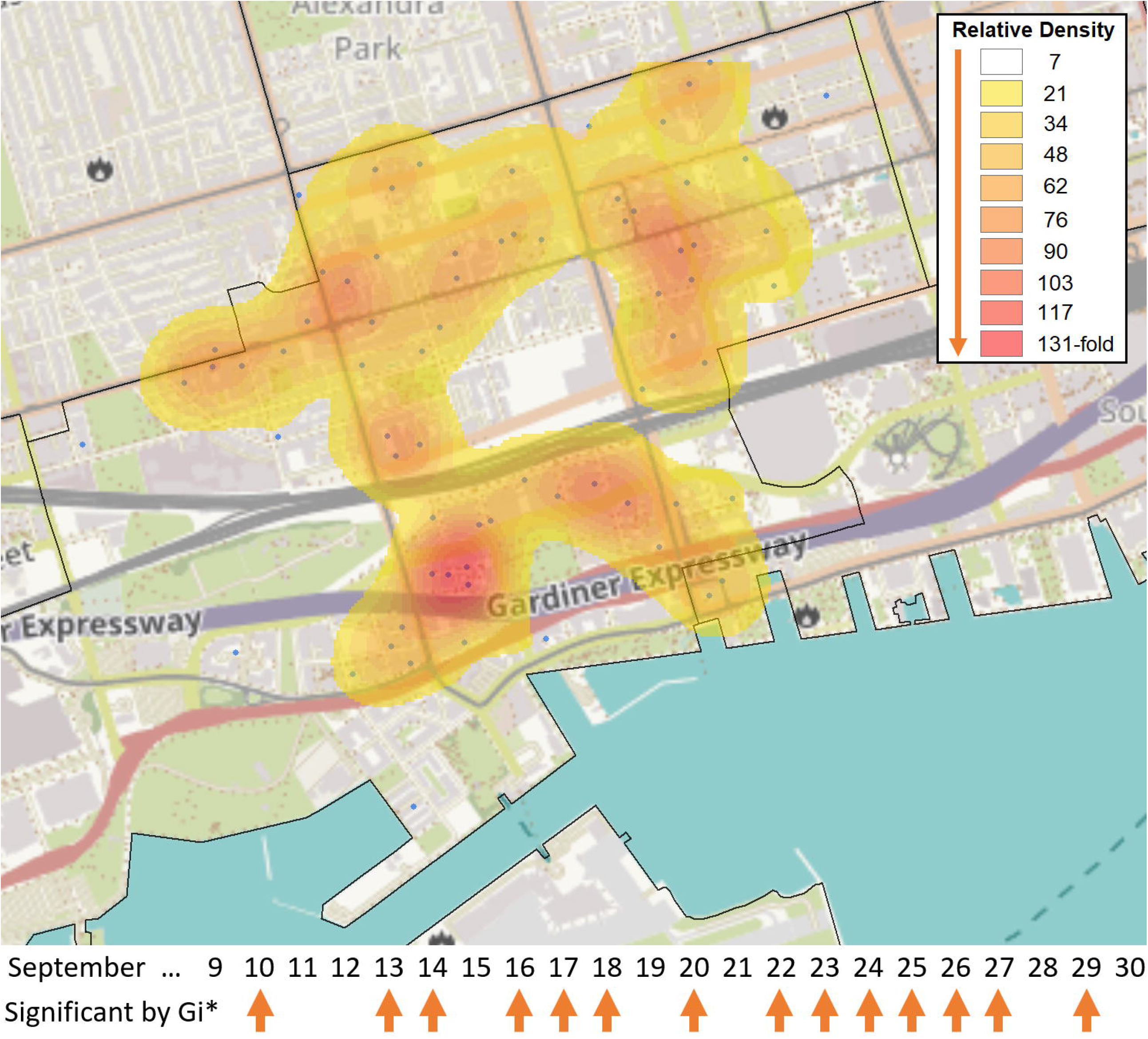
Broad Distribution of COVID-19 Cases across FSA M5V in Toronto. The FSA M5V in the entertainment district of Toronto was identified as a hotspot relative to the rest of Ontario by the Gi* test over 14 days in September 2020 (case counts ≥ 6, with five days exhibiting at least 10 cases [Sept. 10^th^, 13^th^, 20^th^, 22^nd^, 23^rd^]). These cases were distributed across 71 separate PCs in M5V. A heat map generated with the *Kernel Density* function within the Spatial Analyst Toolbox of ArcGIS shows the relative density of postal codes with cases (blue dots), weighted by the number of days in which each individual postal code had ≥1 case (N=118 cases total). Kriging analysis did not identify localized hotspots during this period, as none of the PCs exceeded 3 cases.

Significant FSA hotspots were assessed by Gi* through comparison of cases within KNNs, a fixed distance, or by weighted inverse distance. During the first wave of COVID-19, when testing was not fully established across the province, FSAs with the minimum number of cases considered significant hotspots (≥10) were less common, and fewer FSAs met these criteria (78 to 94%; Extended data^29^, Section 1 - Table S2). Later, fixed distance comparisons identified more FSAs with significantly higher disease burden than other metrics (53% of all FSAs in wave 3 versus 24% for KNN3 and versus 13% with inversely weighted FSAs; Extended data^29^, Section 1 - Table S2). We prioritized the standard Gi* analysis which utilized KNN (where K=3) in this study, as it is moderately stringent (compared to results using the fixed and inverse distance metrics) and is unaffected by the shape of FSAs (which can be quite variable when comparing rural and urban areas).

Space-time Gi* analysis consistently called FSAs with ≥ 10 cases as significant in waves 2 and 3 (>91% and 95%, respectively; Extended data^29^, Section 1 - Table S2). This approach identified nearly twice as many FSAs as significant versus to standard Gi* comparing neighbors up to the same fixed distance (Extended data^29^, Section 1 - Table S2). FSAs deemed significant hotspots by space-time Gi* were tested against many more neighbors (<10 km) relative to FSAs that were not hotspots (averaging 59 and 17 neighbors, respectively). This revealed a bias in significant FSA hotspots towards smaller, densely organized FSAs (which were typically urban), while those that did not achieve significance were often rural, covering larger areas and exhibiting lower COVID-19 counts. Additionally, 98% of the FSAs flagged were also significant hotspots on the days prior and subsequent to the central date in each window. This was uncommon in FSAs that were not determined to be significant hotspots (6%). Temporal trends in the distribution of COVID-19 cases and potential transmission were also identified by Cluster and Outlier analysis of PCs that persisted across a consecutive series of dates (referred to as high-case cluster ‘streaks’; described later in the Results section).

Comparison of kriging and Cluster and Outlier analysis of PC-level case data revealed that the same statistically significant PCs were often identified by both approaches on the same dates (March 26^th^ to December 28^th^, 2020; ≥ 6 cases). A PC was considered significant by Cluster and Outlier analysis if the location was deemed a high-case cluster or outlier with an FDR-correction with 95% confidence, or if ≥5 cases were interpolated within it by kriging. This comparison finds that both approaches are consistent in municipalities with lower overall case counts such as Hamilton (65.2% concordance; N=66 PCs found significant by cluster/outlier analysis), Kitchener / Waterloo (85.1% concordance; N=47), London (69.0% concordance; N=29), and Windsor/Essex (85.0% concordance; N=160) (Extended data^29^, Section 1 - Table S3). The two methods showed <50% correlation to each other when evaluating cases in Ottawa (45.0% concordance; N=211) and Toronto (49.7% concordance; N=529). Nearly all discordant PCs were interpolated to have between 1-5 cases (i.e., marginally discordant). Kriging analysis commonly identified high-case outliers more often than high-case clusters. In Hamilton, 70% of high-case outliers but only 29% of high-case clusters were also found significant by kriging (N=59 and 7, respectively). Similar patterns were observed for all regions evaluated except Windsor/Essex, where overlap between kriging and high-case clusters and outliers were comparable (83% and 96% [N=138 and 22], respectively; Extended data^29^, Section 1 - Table S3).

As incidence rates of COVID-19 have fluctuated since its initial appearance in Canada, the following geostatistical analyses have been stratified by periods with significant infectious burden, which provides a common frame of reference. We organize the analyses according to the three periods (or waves) with maximal occurrence of COVID-19 infections, where wave 1 extended from Mar. 26^th^ to Jun. 16^th^, 2020, wave 2 from Sept. 25^th^, 2020 to Feb. 14^th^, 2021, and wave 3 from Mar. 17^th^ to Jun. 23^rd^, 2021. The intervening periods between waves 1 and 2 and waves 2 and 3 are designated as inter-waves 1-2 and 2-3, respectively.

### Wave 1

Kriging analysis of COVID-19 cases in this wave identified significant localized hotspots in Hamilton, Kitchener/Waterloo, London, Ottawa, Toronto, and Windsor/Essex (all localized hotspots are identified in Extended data^29^, Section 2). High disease burden FSAs (those flagged by Gi* analysis) were often found to include localized hotspots (>50% of flagged FSAs in cities evaluated; Extended data^29^, Section 1 - Table S4). Localized hotspots (regions within contours exceeding significant interpolated count levels) were most frequently identified within high disease burden FSAs in Ottawa during the first COVID-19 wave (84%; Extended data^29^, Section 1 - Table S4). However, kriging also identified 34 additional localized hotspots within FSAs on dates within this period, suggesting that significant COVID-19 events occur that are missed by Gi* analysis. Due to its high density, kriging analysis of cases in Toronto using the Power semivariogram (used in analysis of all other cities) was found to have poor overlap with Gi* analysis (only 31.6% of significant FSAs contained a localized hotspot). The overlap between kriging and Gi* test results was significantly improved using the TPS semivariogram (increased to 60.9%), and this was therefore used for all subsequent kriging analyses of Toronto. PC clustering was uncommon during this wave (<15% of FSAs flagged by Gi* and/or kriging were clustered). The exception was London where 27% of localized hotspots occurred in clustered FSAs [N=11]), and the majority of cases were often localized to a single PC in other cities that were analyzed (Extended data^29^, Section 1 - Table S4).

COVID-19 outbreaks frequently affected residents of long-term care residences during this wave. Geostatistical analyses confirmed the ability to identify these outbreaks. For example, an outbreak in a nursing home in Toronto in early April 2020 (M8V; Lakeshore Lodge) was identified by kriging, while the FSA in which it occurred was also significant by Gi* analysis (Figure 5A). A different outbreak in Toronto in April 2020 (M2K; Extendicare Bayview nursing home) was also detected by kriging analysis, but the FSA in which it was found was not a significant hotspot by Gi* (Figure 5B; it was significant only using a fixed distance metric of ≥ 20km). Similarly, the FSA K1J (Ottawa, ON) was only significant by Gi* using an inverse distance weighted test, despite a mid-April 2020 outbreak in the Rothwell Heights assisted living community which was identified by kriging (Figure 5C). In each of these instances, the interpolation of cases by kriging were similar to actual case counts in these FSAs. This was consistent with the concentration of cases within a single PC (a frequent occurrence in the first COVID- 19 wave; Extended data^29^, Section 1 - Table S4).

**Figure 5.**
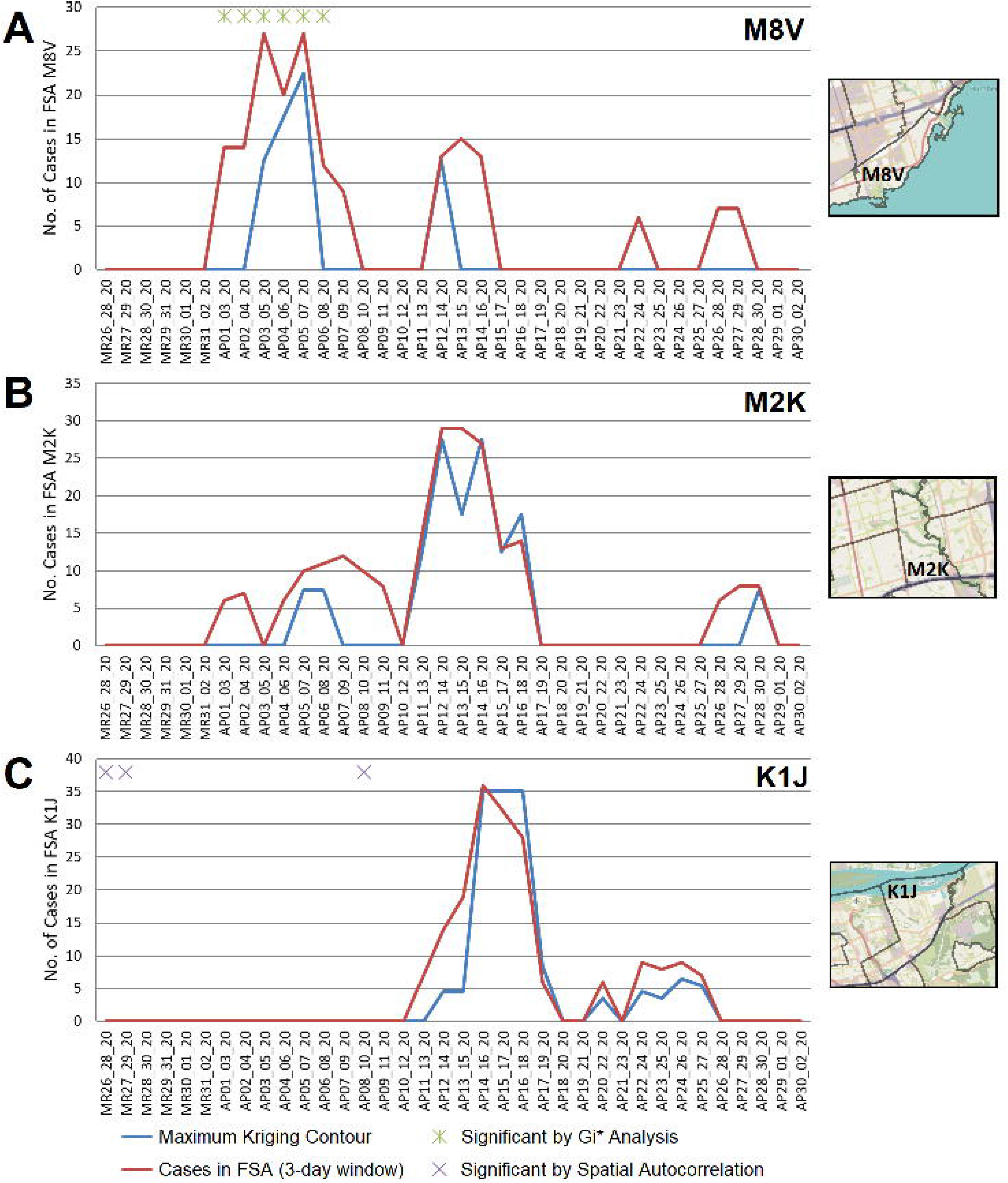
Geostatistical Analysis of COVID-19 Outbreaks during wave 1. FSAs exhibiting high-disease burden by Gi* analysis were identified with localized hotspots by kriging or as having significant case clusters by Spatial Autocorrelation. Panels indicate cases in all PCs within FSAs (A) M8V (B) M2K, and (C) K1J, which were amalgamated over 3-day sliding windows and masked (red line indicates actual case counts; privacy consideration require dates with fewer than 5 cases to be masked as zero). Kriging interpolated the number of cases over these FSAs (blue line), which closely approximates true case counts when counts are high. Green asterisks indicate dates where at least one day within a 3-day window was significant by Gi*; purple crosses indicate significantly clustered cases within PCs.

Localized hotspots identified by kriging comprise a narrow range of PCs in each FSA. The area of interpolated contours that overlapped FSAs with significant disease burdens (by Gi* analysis) was compared to the total area of all corresponding FSAs. COVID-19 cases were highly localized, as the highest value contour (where kriging interpolates the greatest number of cases) constituted only a small portion of the total FSA area (Extended data^29^, Section 1 - Figure S3). While hotspots found by kriging often coincide temporally with Gi* results (Extended data^29^, Section 1 - Table S4), the interpolated hotspots contain fewer cases relative to the entire FSA, since the maxima of kriging contours cover only a portion of the overlapping FSA. The overall area indicated as significant by kriging overlaps a greater fraction of the FSA area, but still covers a fraction of its area (<10% in the majority of cases). Compared to the other COVID-19 waves, however, kriging analysis of cases during this wave tended to span a larger area of each FSA (Extended data^29^, Section 1 - Figure S3A). This appears to be consistent with higher transmissibility (which, we speculate, may be related to lower levels of immunity during this period).

COVID-19 case counts were stratified into 3 categories: biological sex (males vs. females), age (exceeding vs. below 65 years old), and overall health (presence or absence of at-risk, pre-existing comorbidities). Stratified PC-level case counts were evaluated by Cluster and Outlier analysis across all waves (Tables 1A and 1B). In the first wave, the frequency of high-case outliers (a high-case PC with few cases amongst its immediate neighbors) is biased towards older, at-risk populations (Table 1A). Across all municipalities, PCs were flagged nearly 5-fold more often in the older population (305 and 62 PCs were identified as a high-case outlier on different dates in this wave, respectively). A similar pattern was observed in at-risk vs healthy populations (321 and 31 high-case outlier PCs, respectively), which is not surprising due to the significant overlap between the elevated age and groups with at-risk comorbid diagnoses. High-case clusters were identified less often, but also showed this pattern, which is likely related to the high frequency of outbreaks in long-term care homes during this period (Table 1B). Windsor/Essex was an exception to this trend, since the frequencies of PCs designated as high-case outliers were comparable among both age and comorbidity groups (20 vs 17 for older vs younger populations, 19 and 17 for at-risk vs. healthy populations; Table 1A). The frequency of high-case clusters/outliers identified by sex-stratification varied by the region analyzed (e.g., more high-case outliers were identified in the female population in Toronto, in contrast within the male population in Windsor/Essex).

**Table 1.** A. Distribution of High-Case *Outlier Postal Codes* in Different Municipalities

| City | Wave<sup>1</sup> | Number of High-Case *Outlier* PCs in Stratified Populations<sup>2</sup> |  |  |  |  |  |  |
| --- | --- | --- | --- | --- | --- | --- | --- | --- |
|  |  | All Cases | Males | Females | ≥65 years | <65 years | At-Risk | Healthy |
| Hamilton | 1 | 8 | 3 | 6 | 7 | 1 | 8 | 1 |
|  | 1-2 | 0 | 0 | 0 | 0 | 0 | 0 | 0 |
|  | 2 | 100 | 20 | 43 | 31 | 45 | 41 | 24 |
|  | 2-3 | 29 | 6 | 4 | 0 | 25 | 2 | 8 |
|  | 3 | 150 | 36 | 43 | 0 | 131 | 9 | 84 |
| Kitchener /<br>Waterloo | 1 | 25 | 14 | 11 | 17 | 4 | 24 | 0 |
|  | 1-2 | 0 | 0 | 0 | 0 | 0 | 0 | 0 |
|  | 2 | 31 | 3 | 10 | 12 | 13 | 17 | 6 |
|  | 2-3 | 11 | 5 | 0 | 0 | 11 | 0 | 8 |
|  | 3 | 30 | 3 | 0 | 0 | 26 | 0 | 15 |
| London | 1 | 11 | 3 | 3 | 6 | 5 | 6 | 3 |
|  | 1-2 | 6 | 0 | 2 | 0 | 6 | 0 | 0 |
|  | 2 | 31 | 3 | 5 | 4 | 21 | 5 | 7 |
|  | 2-3 | 2 | 0 | 0 | 0 | 0 | 2 | 0 |
|  | 3 | 28 | 6 | 1 | 0 | 26 | 0 | 10 |
| Ottawa | 1 | 50 | 10 | 26 | 42 | 2 | 45 | 0 |
|  | 1-2 | 28 | 2 | 10 | 8 | 16 | 11 | 12 |
|  | 2 | 120 | 25 | 21 | 28 | 84 | 29 | 54 |
|  | 2-3 | 12 | 0 | 2 | 2 | 7 | 2 | 7 |
|  | 3 | 50 | 8 | 13 | 0 | 48 | 0 | 30 |
| Toronto | 1 | 261 | 81 | 162 | 213 | 33 | 219 | 10 |
|  | 1-2 | 25 | 3 | 10 | 6 | 13 | 10 | 4 |
|  | 2 | 596 | 78 | 178 | 191 | 315 | 216 | 127 |
|  | 2-3 | 89 | 17 | 13 | 1 | 76 | 16 | 40 |
|  | 3 | 428 | 45 | 72 | 3 | 357 | 25 | 181 |
| Windsor /<br>Essex | 1 | 42 | 25 | 13 | 20 | 17 | 19 | 17 |
|  | 1-2 | 35 | 27 | 9 | 3 | 34 | 3 | 30 |
|  | 2 | 212 | 83 | 91 | 86 | 121 | 116 | 68 |
|  | 2-3 | 6 | 3 | 0 | 0 | 3 | 3 | 3 |
|  | 3 | 43 | 11 | 8 | 0 | 41 | 2 | 31 |

<sup>1</sup> Interwaves 1-2 and 2-3 are displayed as '1-2' and '2-3', respectively.
<sup>2</sup> PCs identified as a high-case outlier on a particular 3-day interval with ≥ 6 cases when analyzing stratified case data.

B. Distribution of High-Case *Cluster Postal Codes* in Different Municipalities
| City | Wave<sup>1</sup> | Number of High-Case *Clustered* PCs in Stratified Populations<sup>2</sup> |  |  |  |  |  |  |
| --- | --- | --- | --- | --- | --- | --- | --- | --- |
|  |  | All Cases | Males | Females | ≥65 years | <65 years | At-Risk | Healthy |
| Hamilton | 1 | 2 | 0 | 0 | 1 | 0 | 1 | 0 |
|  | 1-2 | 0 | 0 | 0 | 0 | 0 | 0 | 0 |
|  | 2 | 28 | 5 | 4 | 0 | 23 | 0 | 10 |
|  | 2-3 | 5 | 0 | 1 | 0 | 5 | 0 | 1 |
|  | 3 | 52 | 25 | 17 | 0 | 47 | 5 | 34 |
| Kitchener /<br>Waterloo | 1 | 3 | 0 | 3 | 3 | 0 | 3 | 0 |
|  | 1-2 | 0 | 0 | 0 | 0 | 0 | 0 | 0 |
|  | 2 | 3 | 1 | 0 | 0 | 1 | 0 | 0 |
|  | 2-3 | 2 | 0 | 0 | 0 | 2 | 0 | 0 |
|  | 3 | 1 | 0 | 0 | 0 | 0 | 0 | 0 |
| London | 1 | 0 | 0 | 0 | 0 | 0 | 0 | 0 |
|  | 1-2 | 0 | 0 | 0 | 0 | 0 | 0 | 0 |
|  | 2 | 9 | 0 | 1 | 1 | 4 | 4 | 0 |
|  | 2-3 | 0 | 0 | 0 | 0 | 0 | 0 | 0 |
|  | 3 | 7 | 0 | 0 | 0 | 7 | 0 | 3 |
| Ottawa | 1 | 8 | 1 | 0 | 6 | 0 | 7 | 0 |
|  | 1-2 | 5 | 0 | 1 | 0 | 5 | 0 | 3 |
|  | 2 | 21 | 2 | 4 | 0 | 17 | 0 | 7 |
|  | 2-3 | 1 | 0 | 0 | 0 | 1 | 0 | 0 |
|  | 3 | 28 | 6 | 7 | 0 | 22 | 0 | 17 |
| Toronto | 1 | 22 | 0 | 7 | 2 | 8 | 9 | 1 |
|  | 1-2 | 4 | 0 | 4 | 4 | 0 | 4 | 0 |
|  | 2 | 183 | 19 | 30 | 7 | 149 | 11 | 73 |
|  | 2-3 | 72 | 9 | 9 | 1 | 57 | 6 | 19 |
|  | 3 | 172 | 11 | 12 | 0 | 136 | 3 | 51 |
| Windsor /<br>Essex | 1 | 2 | 2 | 0 | 0 | 2 | 0 | 2 |
|  | 1-2 | 17 | 7 | 1 | 0 | 15 | 0 | 12 |
|  | 2 | 36 | 13 | 4 | 0 | 23 | 0 | 11 |
|  | 2-3 | 1 | 0 | 0 | 0 | 1 | 0 | 0 |
|  | 3 | 5 | 0 | 0 | 0 | 5 | 0 | 0 |

<sup>1</sup> Interwaves 1-2 and 2-3 are displayed as '1-2' and '2-3', respectively.
<sup>2</sup> PCs identified as a high-case cluster on a particular 3-day interval with ≥ 6 cases when analyzing stratified case data.

### Interwave 1-2

The first COVID-19 wave ended approximately two months after the imposition of public health prohibitions on mobility, limits on assembly, and social distancing (i.e., a lockdown) in Ontario. June 16^th^, 2020 defined the start of interwave 1-2, a period of much lower COVID-19 incidence than the preceding 2 1/2 months. Fewer FSAs exhibited high disease burden by Gi* analysis (39.7% of those called in wave 1, despite the duration of interwave 1-2 being longer; Extended data^29^, Section 1 - Table S2). Despite the overall decrease in cases province-wide, the frequency of FSAs in the Windsor/Essex region that were designated hotspots by both Gi* and kriging analysis increased relative to the preceding wave (Extended data^29^, Section 1 - Table S4). This was due to multiple outbreaks in the Essex County FSAs in Leamington [N8H] and Kingsville [N9Y] (Extended data^29^, Section 1 - Figure S4). These outbreaks were publicly documented in guest farm workers, which is reflected by the Cluster and Outlier analysis of stratified case data (a strong bias of high-case clusters and outliers in healthy males < 65 years in Windsor/Essex; Extended data^29^, Section 1 - Table S4). These same locations were correlated with interpolated kriging contour maxima and often consisted of isolated single PCs (Tables 1A and 1B). However, there were instances where the interpolated count values exceeded the actual case count (i.e., N8H, June 22-26^th^, 2020; Extended data^29^, Section 1 - Figure S4B) due to concurrent high case counts from neighboring FSAs (i.e., N0P 2J0; Extended data^29^, Section 1 - Figure S4C) bleeding into these contours from a single PC.

### Wave 2

FSAs with high disease burden were more frequent within the second wave (Extended data^29^, Section 1 - Table S2). Even when accounting for its longer duration, there was a >10-fold increase in FSAs with high disease burden based on the Gi* test. The distribution of cases was more widespread across each FSA, as COVID-19 testing became more reliable and pervasive. Localized hotspots were detected less frequently within FSAs with high disease burden, i.e., significant FSAs by Gi* analysis overlapped less often with significant kriging contours in all municipalities except Windsor/Essex (Extended data^29^, Section 1 - Table S4). Additionally, interpolated case counts by kriging tended to be less than actual FSA case counts, due to dispersion of cases across more PCs. Most municipalities contained more FSAs with localized hotspots that were not significant by Gi* analysis. Increased case counts across the province during this wave increased the significance threshold for Gi* analysis. This decreased the likelihood that FSAs in municipalities with moderate population densities were designated significant hotspots. For example, FSAs within Kitchener/Waterloo and London were not significant COVID-19 hotspots during wave 2 (Extended data^29^, Section 1 - Table S4). Hotspots in these regions were coincident with maximum contour levels interpolated by kriging. PCs had a greater proportion of high-case clusters and/or outliers in younger and healthy populations, in contrast with wave 1 (Tables 1A and 1B). The reduction of COVID-19 outbreaks (i.e., hotspots) in long-term care homes during this period was supported by reduced statistical significance of counts at these locations relative to increased background case counts across the province.

Localized hotspots detected in PCs by kriging were more frequent in Hamilton, London, Ottawa, Toronto and Windsor/Essex during this wave than in the preceding one (Extended data^29^, Section 1 - Table S4). For example, an interpolated hotspot in the N5Y FSA (London) with significant cases over consecutive 3-day intervals (December 19-21 and December 20-22, 2020; Figure 6) represents a ‘streak’, since kriging analysis interpolated cases over this FSA throughout the previous week (however, it was below the kriging threshold for significance). This hotspot encompasses a 2 building apartment complex that was publicized on December 29 by the local health authorities as a COVID-19 hotspot, two weeks after it was first detected by kriging^32^. While the interpolated case count is lower than the total count in this FSA on this date (N=33, other cases are present at other locations), this hotspot encompasses only 4% of the FSA by area. Other localized hotspots during this wave are depicted (e.g. in Leamington ON; Extended data^29^, Section 1 - Figure S5) and catalogued in Extended data^29^, Section 2.

**Figure 6.**
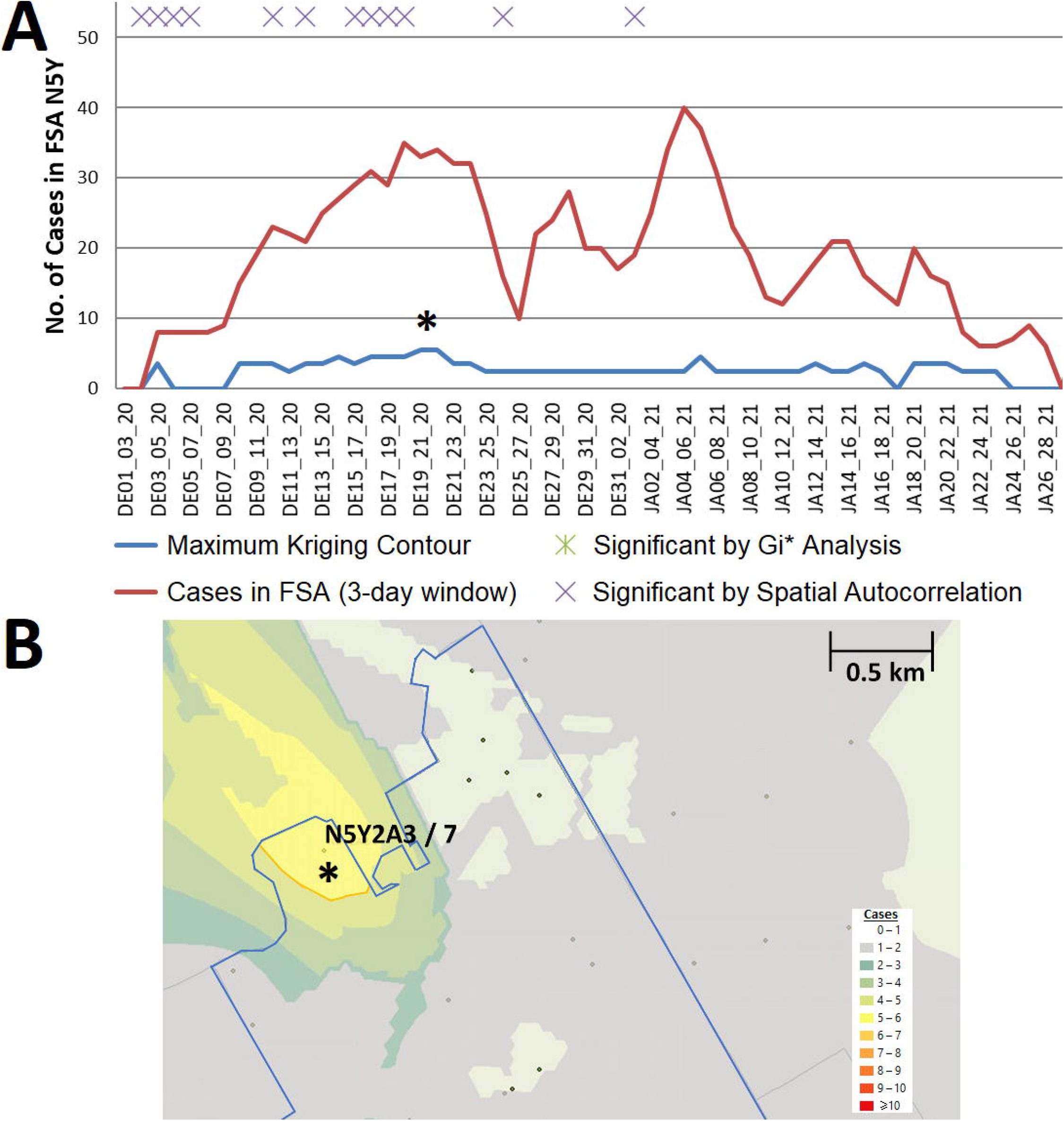
Example of a Localized Hotspot in London Ontario. (A) Geostatistical analysis of the FSA N5Y (London, ON) identifies a significant localized hotspot from Dec. 19^th^ to the 22^nd^. Although significant, estimates from kriging were lower than actual case counts. B) The kriging contour of a localized hotspot (yellow) interpolated with ≥5 cases encompasses 4% of the FSA area. Dots indicate PCs at least 1 case, however only those with ≥6 cases are labelled (with PC identifier / case count).

Significant clustering of cases in PCs in close proximity based on Spatial Autocorrelation analysis may be distinct from other tests that identify localized hotspots. Spatially clustered localized COVID-19 hotspots may not be confined within the same region of an FSA. For example, FSA M1P (Toronto) from Nov. 1-3 to Nov. 3-5, 2020 exhibited significant case clustering based on Gi* analysis. However, a localized hotspot identified within this FSA during this time did not overlap this cluster (Extended data^29^, Section 1 - Figure S6A). A neighboring FSA M1C showed significant spatially correlated cases within the same timeframe (Nov. 6-8, 2020), however localized hotspots were not detected in this FSA (Extended data^29^, Section 1 - Figure S6B). No PC within this FSA had ≥6 cases during this period. This illustrates how combining Spatial Autocorrelation, Gi* and kriging analyses can improve the interpretation of asymmetric COVID-19 case distributions.

### Interwave 2-3

The interval between the end of wave 2 and inception of wave 3 was short (∼31 days), however by comparison with interwave 1-2, the case counts were considerably higher (averaging 1134 and 154 per day, respectively). Thus, a far greater number of high disease burden FSAs were identified during this period (1.9 and 4.7-fold more often compared to wave 1 and interwave 1-2, respectively; Extended data^29^, Section 1 - Table S2). However, most of these occurred in Toronto and Windsor/Essex regions only (Extended data^29^, Section 1 - Table S4).

### Wave 3

The frequencies of high disease burden FSAs in waves 2 and 3 were comparable upon adjusting for duration of each wave (Extended data^29^, Section 1 - Table S2). The regions detected by geostatistical analyses were different. High disease burden FSAs implicated in Hamilton and Kitchener/Waterloo by Gi* analysis were significantly more frequent compared to the preceding wave, while FSAs within Toronto were less often significant (Extended data^29^, Section 1 - Table S4). By contrast, localized hotspots within FSAs were generally less frequent than in wave 2, most notably in the Windsor/Essex region which contained 389 localized hotspots in wave 2, but 80 in wave 3 (Extended data^29^, Section 1 - Table S4).

High-case clusters and outliers were seldomly identified in older (≥65) populations in wave 3 and interwave 2-3 (Tables 1A and 1B). Similarly, there is a considerable decrease in the number of case clusters/outliers in at-risk populations relative to previous waves. For example, analysis of at-risk populations in Windsor/Essex identified 116 PCs as a high case outlier on a particular date during wave 2, but only 2 in wave 3 (Table 1A). These observations may be related to effectiveness of vaccination, based on the initial prioritization of older and at-risk cohorts for immunization.

### The distribution of and relationships between clustered PCs with persistent COVID-19 cases

COVID-19 distributions were evaluated by Cluster and Outlier analysis to identify neighboring PCs with high concentrations of cases. PCs deemed part of a high-case cluster that were clustered over contiguous date ranges were then identified. These PCs were recognized as clustered over the duration of a time “streak”, which by definition, allows for a single day in which the PC was either not clustered or did not meet the minimum case count threshold of ≥ 6 cases within the 3-day sliding window. PCs that were part of a high-case cluster, but did not meet the case threshold, were not reported. This analysis identified 20 unique PCs with one or more high-case cluster streaks in Hamilton, 5 in Kitchener/Waterloo, 10 in London, 24 in Ottawa, 216 in Toronto, and 19 in Windsor/Essex (Extended data^29^, Section 1 - Table S5). Streaks were uncommon in Kitchener/Waterloo and London, as PCs in these cities rarely fulfilled the minimum case threshold. Most streaks were shorter than 4 sliding window periods, indicating that the most frequent streaks were brief due the large number of single day discontinuities from case reporting patterns and ascertainment bias. For these reasons, ≤ 3-day bins were not considered evidence of a streak. However, 37 streaks longer than 3 days were interpreted to indicate persistent infections at the same locations. Streaks with more than 3 consecutive high-case clusters were uncommon in wave 1, with M6M 2J5 (West Park Long Term Care Center) as the only example of a streak exceeding 3 days. The lack of other locations with persistent infections may be related to inconsistent case reporting which was hampered by limited testing during this period^4,33^.

We sought evidence of COVID-19 transmission by identifying pairs of PCs with high case counts that were adjacent to one another by both distance and time. In Figure 7A, we analyzed PCs and their 10 closest neighbors, where streaks were either concurrent, consecutive, or separated by short gaps within the same wave. Concurrent streaks of neighboring PC pairs spanned at least 3 and as many as 29 days in Hamilton, Ottawa, Toronto, and Windsor/Essex (Table 2). There were a greater number of discontinuous streaks across all municipalities, with gaps separating streaks in neighboring PCs varied from short to long intervals (Extended data^29^, Section 1 - Table S6). During the second wave, many pairs of PCs were comprised of neighboring apartment buildings, for example, within Toronto FSA M4H. Other long concurrent paired streaks (exceeding 1 week in duration) occurred in Toronto PCs M1R 1S9 and M1R 1T1, M1L 1K9 and M1L 1L1 (both PC pairs in North York, Toronto), and L0R 1C0 with its neighbors in Hamilton (Table 2). Pairs of concurrent streaks in close proximity did not occur during wave 1, since streaks were uncommon during this time. However, many neighboring PCs were both found to be clustered both spatially and temporally during interwave 1-2 and waves 2 and 3 (Extended data^29^, Section 1 - Table S6). While not concurrent, neighboring PC pairs in rural Essex county were clustered within 10 days of one another during interwave 1-2 (N0P 2L0, N0P 2P0 and N0R 1B0). This is confirmed by publicly reported outbreaks in agriculture facilities of this region during this time. During wave 2, 56 unique pairs of streaks were identified in neighboring PCs in Toronto (18 clustered concurrently, another 14 within a week of one other), 4 pairs in Hamilton (2 concurrent), 1 in London (concurrent), 4 in Ottawa (1 concurrent) and 14 in Windsor/Essex (10 concurrent; all involve PCs in Essex county). There were fewer incidences of streaks during the third wave, which may be related to the effectiveness of vaccination efforts which limit transmission (Figure 7B). During wave 3, we identified an additional 30 pairs of PCs with streaks in Toronto (7 are clustered concurrently, and an additional 4 pairs are offset by 1 week), and 6 in Hamilton (3 concurrent, and another pair offset by 2 days). Interestingly, 9 of the same PC pairs in Toronto clustered during the third wave were also flagged during the second wave, which could be an indication of an undetected reservoir of COVID-19 that persisted in this population during interwave 2-3. These pairs of PCs occurred in the Toronto neighbourhoods of East York (M4H 1J4 and M4H 1J5), Scarborough (M1H 2Y7 and M1H 2E9), North York (M1R 1T1 and M1R 1S9; M6M 5B3 and M6M 5B7), and Etobicoke (M9C 1G7 and M9C 1G6; M9V 3S6 and M9V 4A4; M9V 3S6 and M9V 4M1; M9V 4A4 and M9V 4M1; M9W 6L4 and M9W 6A7).

**Table 2.**
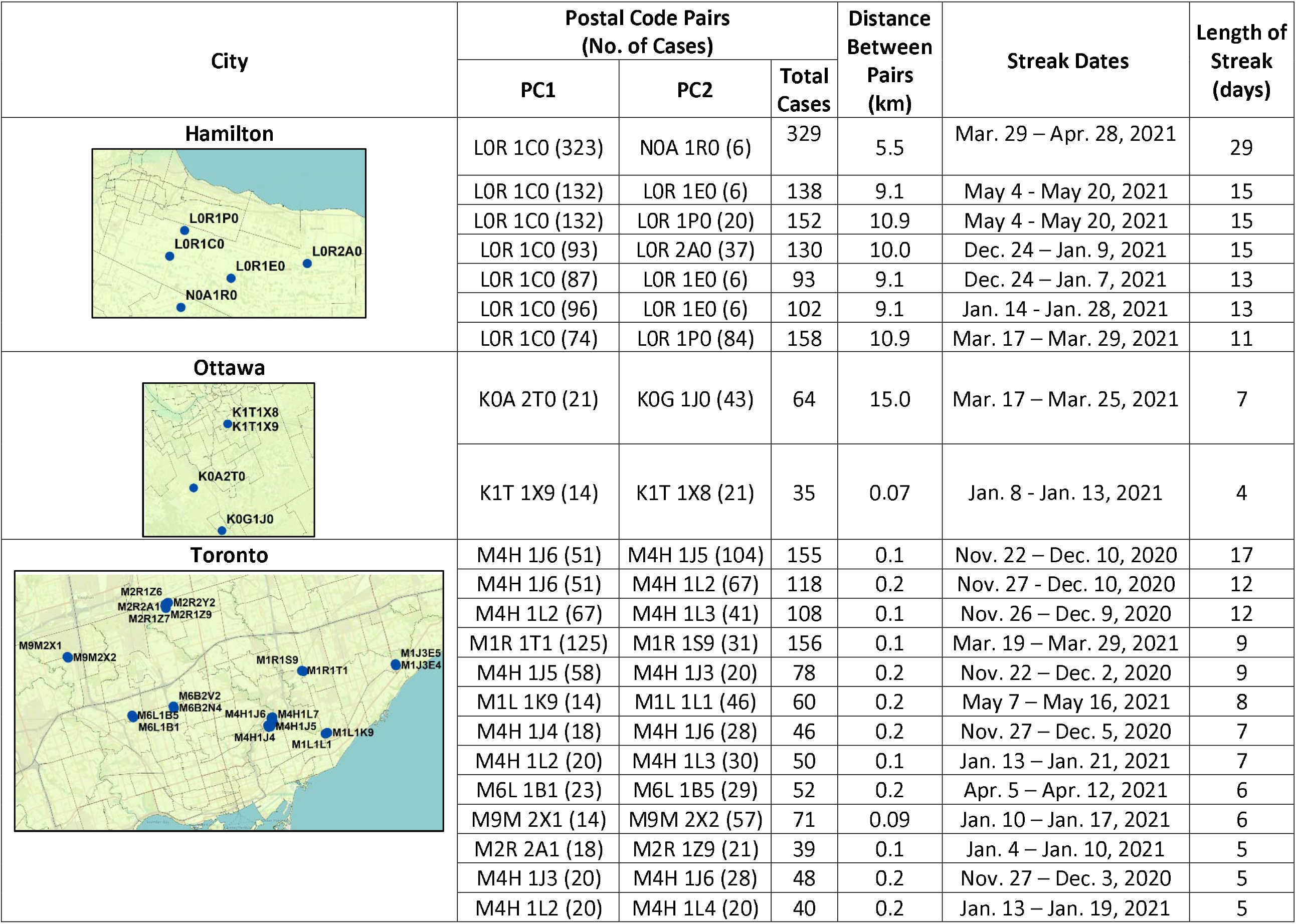
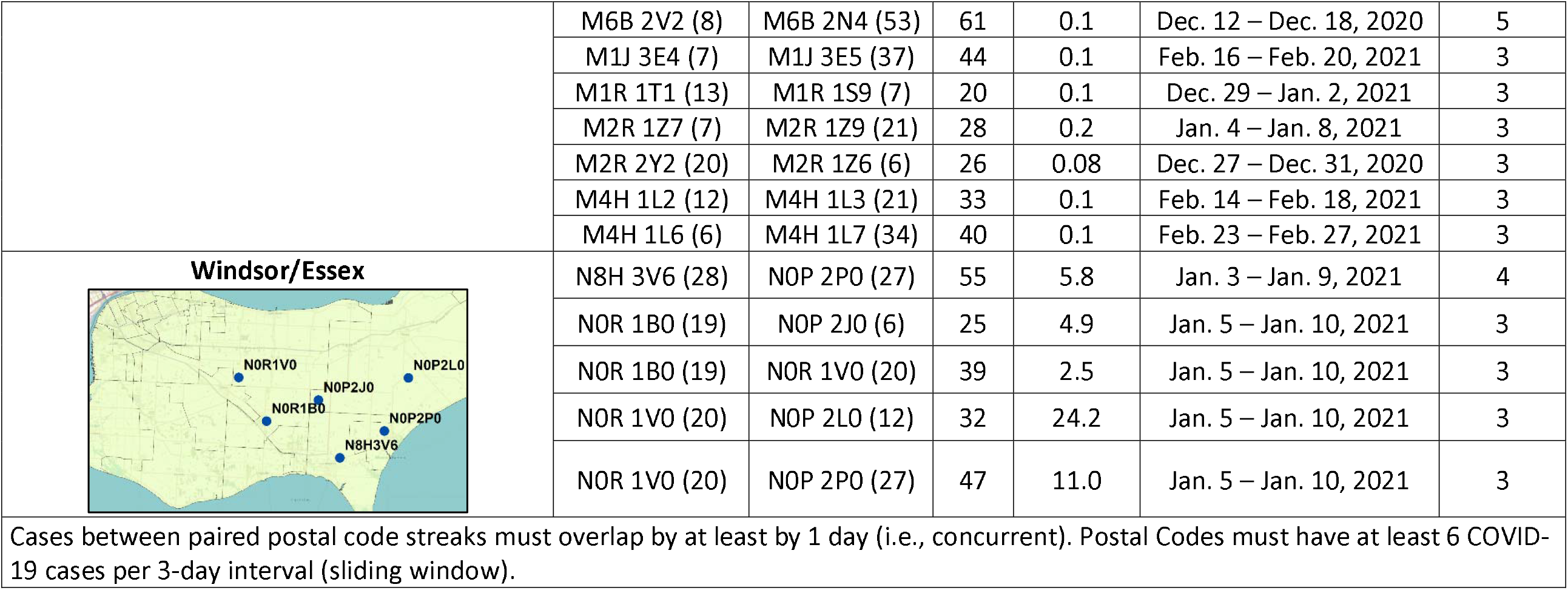
Longest Concurrent Streaks between Postal Code Neighbours.

| City | Postal Code Pairs<br>(No. of Cases) |  |  | Distance<br>Between<br>Pairs<br>(km) | Streak Dates | Length of<br>Streak<br>(days) |
| --- | --- | --- | --- | --- | --- | --- |
|  | PC1 | PC2 | Total<br>Cases |  |  |  |
| **Hamilton** | L0R 1C0 (323) | N0A 1R0 (6) | 329 | 5.5 | Mar. 29 – Apr. 28, 2021 | 29 |
|  | L0R 1C0 (132) | L0R 1E0 (6) | 138 | 9.1 | May 4 - May 20, 2021 | 15 |
|  | L0R 1C0 (132) | L0R 1P0 (20) | 152 | 10.9 | May 4 - May 20, 2021 | 15 |
|  | L0R 1C0 (93) | L0R 2A0 (37) | 130 | 10.0 | Dec. 24 – Jan. 9, 2021 | 15 |
|  | L0R 1C0 (87) | L0R 1E0 (6) | 93 | 9.1 | Dec. 24 – Jan. 7, 2021 | 13 |
|  | L0R 1C0 (96) | L0R 1E0 (6) | 102 | 9.1 | Jan. 14 - Jan. 28, 2021 | 13 |
|  | L0R 1C0 (74) | L0R 1P0 (84) | 158 | 10.9 | Mar. 17 – Mar. 29, 2021 | 11 |
| **Ottawa** | K0A 2T0 (21) | K0G 1J0 (43) | 64 | 15.0 | Mar. 17 – Mar. 25, 2021 | 7 |
|  | K1T 1X9 (14) | K1T 1X8 (21) | 35 | 0.07 | Jan. 8 - Jan. 13, 2021 | 4 |
| **Toronto** | M4H 1J6 (51) | M4H 1J5 (104) | 155 | 0.1 | Nov. 22 – Dec. 10, 2020 | 17 |
|  | M4H 1J6 (51) | M4H 1L2 (67) | 118 | 0.2 | Nov. 27 - Dec. 10, 2020 | 12 |
|  | M4H 1L2 (67) | M4H 1L3 (41) | 108 | 0.1 | Nov. 26 – Dec. 9, 2020 | 12 |
|  | M1R 1T1 (125) | M1R 1S9 (31) | 156 | 0.1 | Mar. 19 – Mar. 29, 2021 | 9 |
|  | M4H 1J5 (58) | M4H 1J3 (20) | 78 | 0.2 | Nov. 22 – Dec. 2, 2020 | 9 |
|  | M1L 1K9 (14) | M1L 1L1 (46) | 60 | 0.2 | May 7 – May 16, 2021 | 8 |
|  | M4H 1J4 (18) | M4H 1J6 (28) | 46 | 0.2 | Nov. 27 – Dec. 5, 2020 | 7 |
|  | M4H 1L2 (20) | M4H 1L3 (30) | 50 | 0.1 | Jan. 13 – Jan. 21, 2021 | 7 |
|  | M6L 1B1 (23) | M6L 1B5 (29) | 52 | 0.2 | Apr. 5 – Apr. 12, 2021 | 6 |
|  | M9M 2X1 (14) | M9M 2X2 (57) | 71 | 0.09 | Jan. 10 – Jan. 17, 2021 | 6 |
|  | M2R 2A1 (18) | M2R 1Z9 (21) | 39 | 0.1 | Jan. 4 – Jan. 10, 2021 | 5 |
|  | M4H 1J3 (20) | M4H 1J6 (28) | 48 | 0.2 | Nov. 27 – Dec. 3, 2020 | 5 |
|  | M4H 1L2 (20) | M4H 1L4 (20) | 40 | 0.2 | Jan. 13 – Jan. 19, 2021 | 5 |
|  | M6B 2V2 (8) | M6B 2N4 (53) | 61 | 0.1 | Dec. 12 – Dec. 18, 2020 | 5 |
|  | M1J 3E4 (7) | M1J 3E5 (37) | 44 | 0.1 | Feb. 16 – Feb. 20, 2021 | 3 |
|  | M1R 1T1 (13) | M1R 1S9 (7) | 20 | 0.1 | Dec. 29 – Jan. 2, 2021 | 3 |
|  | M2R 1Z7 (7) | M2R 1Z9 (21) | 28 | 0.2 | Jan. 4 – Jan. 8, 2021 | 3 |
|  | M2R 2Y2 (20) | M2R 1Z6 (6) | 26 | 0.08 | Dec. 27 – Dec. 31, 2020 | 3 |
|  | M4H 1L2 (12) | M4H 1L3 (21) | 33 | 0.1 | Feb. 14 – Feb. 18, 2021 | 3 |
|  | M4H 1L6 (6) | M4H 1L7 (34) | 40 | 0.1 | Feb. 23 – Feb. 27, 2021 | 3 |
| **Windsor/Essex** | N8H 3V6 (28) | N0P 2P0 (27) | 55 | 5.8 | Jan. 3 – Jan. 9, 2021 | 4 |
|  | N0R 1B0 (19) | N0P 2J0 (6) | 25 | 4.9 | Jan. 5 – Jan. 10, 2021 | 3 |
|  | N0R 1B0 (19) | N0R 1V0 (20) | 39 | 2.5 | Jan. 5 – Jan. 10, 2021 | 3 |
|  | N0R 1V0 (20) | N0P 2L0 (12) | 32 | 24.2 | Jan. 5 – Jan. 10, 2021 | 3 |
|  | N0R 1V0 (20) | N0P 2P0 (27) | 47 | 11.0 | Jan. 5 – Jan. 10, 2021 | 3 |
| Cases between paired postal code streaks must overlap by at least by 1 day (i.e., concurrent). Postal Codes must have at least 6 COVID-19 cases per 3-day interval (sliding window). |  |  |  |  |  |  |

**Figure 7.**
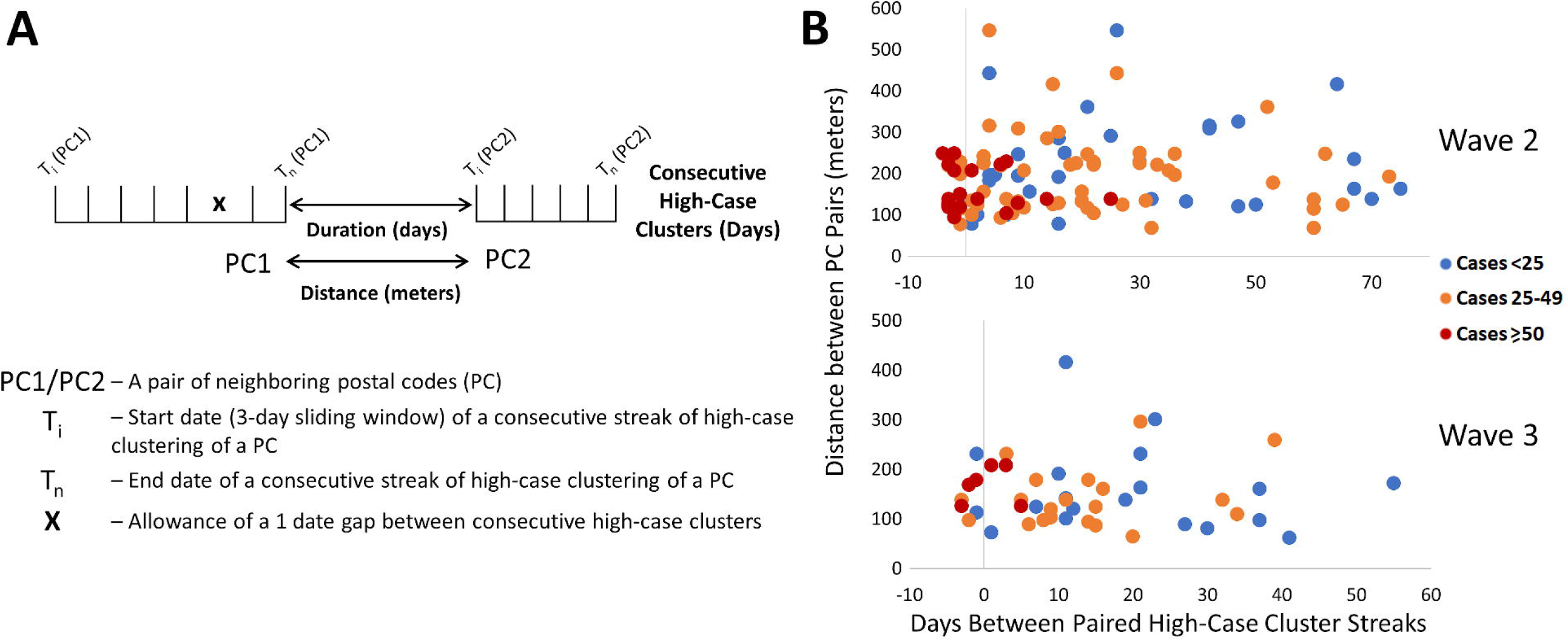
Contiguous or Consecutive Pairs of Toronto Postal Codes with COVID-19 Case Clusters lasting multiple days. Pairs of neighboring PCs with high-case count clusters were identified by Cluster and Outlier analysis. Panels show: (A) parameters measured in PC pairs that were elements of significant high-case clusters over multiple consecutive days (allowing for a single day gap), termed “streaks”; (B) characteristics of neighboring pairs of PCs with multi-day streaks in close proximity in Toronto (of ten closest neighbors based on distance between centroids) during waves 2 (top) and 3 (bottom). Wave 1 is not indicated because streaks were infrequent. Each point represents a pair of PCs, with color codes (see legend) indicating ranges of total number of cases in both, the X axis indicates the duration between PC pairs (with negative values representing number of days of overlap between end of first PC streak and beginning of the second), and the Y axis indicates spatial distance between PCs constituting the pair.

We examined evidence for likely community spread of COVID-19 between infected individuals in different pairs of neighboring PCs. This involved determining the distances separating them, the duration between streaks in the respective PCs, the total case counts among PC pairs during these streaks, and then assessing the significance of these characteristics. During waves 2 and 3 in Toronto, paired concurrent or consecutive streaks in such PCs were common (Figure 7B). Frequencies of these PC pairs were negatively correlated with the duration of the gap between the end of a streak in one member of the pair and the beginning of the other (r=-0.57 and -0.50 for waves 2 and 3, respectively).

To assess the significance of these results, a multinomial logistic regression model was derived from (1) the total number of cases in each pair of PCs over the course of both streaks (classified as either <25, between 25 and 49, or ≥50 cases), (2) the duration or gap (in days) separating streaks in a pair of PCs, and (3) the distances between the centroids of both PCs. The duration between streaks was significantly shorter for PC pairs with ≥50 cases relative to those with <25 cases (p=0.0005 and 0.0141 for waves 2 and 3, respectively, using the Wald two-tailed z-test on the coefficients of the model). The same relationship was also significant when comparing streaks in pairs of PCs with ≥50 cases and those with 25-49 cases (p= 0.0015 and 0.0277 for waves 2 and 3, respectively). Total case counts and time between streaks in a pair of PCs were also inversely correlated (r= -0.41 in both waves 2 and 3), which is consistent with disease transmission between adjacent PCs being more common when cases were more abundant. These parameters were also inversely correlated (r=-0.13 and -0.17, respectively) in Ottawa and Windsor/Essex, but under less stringent criteria that allowed ≥6 cases for only one of the PCs in a pair on a single day. The relationship between distance between PCs and total number of cases was not significant by the regression model in either of these waves. Increasing the number of PC neighbors considered over a larger range of distances did not alter this result (Extended data^29^, Section 1 - Figure S7). However, upon dividing groups of neighboring PCs thresholded according to average quartile values (distance [182m] and average gap between streaks [17.5 days]) distance between neighbors was significant when combined with the degree to which they coincided temporally. Below average duration between streaks and distance separating PCs were 2.8-fold more likely to exhibit ≥ 50 cases than all other PC pairs (p=0.002 by Mantel-Haenszel chi square test). Transmission of higher levels of COVID-19 between pairs of PCs is supported by abbreviated intervals between streaks and their proximity to one another.

Multiple paired PC streaks in Toronto were tightly grouped within a single neighborhood over a short period of time (Extended data^29^, Section 1 - Table S6). Directional network diagrams were created to illustrate the connectivity and temporal relationships between these and other paired high-case cluster streaks in close proximity (Extended data^29^, Section 1 - Figures S8 and S9 for waves 2 and 3, respectively). The preponderance of paired streaks were bimodal (9 paired streaks where only two neighbors interact [Extended data^29^, Section 1 - Figure S8D], of which 17 occurred in wave 3 [Extended data^29^, Section 1 - Figure S9A]), and in close succession (≤7 days between streaks). Multi-node networks involving ≥3 postal codes occurred in 3 distinct neighborhoods in wave 2, and another three neighborhoods within wave 3. These neighborhoods were often comprised of multiple adjacent apartment buildings, and rarely occurred in low-density residential areas. The PCs where streaks tended to coincide were often proximate to facilities where communities may have congregated (grocery stores, schools, parks, or religious sites). We investigated whether the participation of many PCs in a multi-node network was a result of higher case counts during a streak (which would suggest an increased probability of transmission between these locations). However, bimodal networks of two PCs with recurrent high case counts were not rare (e.g., >100 cases within a 9-day streak in M1R 1T1), so other factors are necessary to explain why some PCs form multi-node networks. Furthermore, the average numbers of cases within streaks in bimodal networks and multi-node networks were comparable (18.8 and 17.0 cases per streak, respectively).

In some instances, PCs began to accrue cases prior to their inclusion in significant clusters of PCs or reaching the minimum numbers of cases to be designated a COVID-19 hotspot (M4H 1K1, M4H 1K2, M4H 1K4, M4H 1L1, M4H 1L6, and M4H 1L7). These PCs were nevertheless elements of the M4H FSA streak. Similarly, M4H 1L3 and M4H 1L4 were elements of a cluster containing M4H 1J4 on Nov. 8-10, 2020, at least 2 weeks prior to the occurrence of the corresponding multi-day streaks. Consecutive streaks within a set of clustered PCs in Etobicoke (M9V 3S6, M9V 3Z8, M9V 4A4, M9V 4A9, M9V 4M1, M9V 4P1, and M9V 5G9; centroids were all within 0.63km of one another) occurred between mid-October and late December 2020 (Figure 8B). M9V 5G9 is separated from the other PCs by a wooded area, but all are close to a major thoroughfare, Kipling Avenue. A striking directional acyclic network of COVID-19 cases involved 7 adjacent PCs whose streaks occurred concurrently and were occurred nearly consecutively along Bathurst Street, Toronto (M2R 1Z2, M2R 1Z7, M2R 1Z8, M2R 1Z9, M2R 2A1, M2R 1Z6, M2R 2Y2; within 0.57km of one another; Figure 8C). Curiously, the order of occurrence of COVID-19 infections among these PCs consistently followed a southerly direction along Bathurst Street.

**Figure 8.**
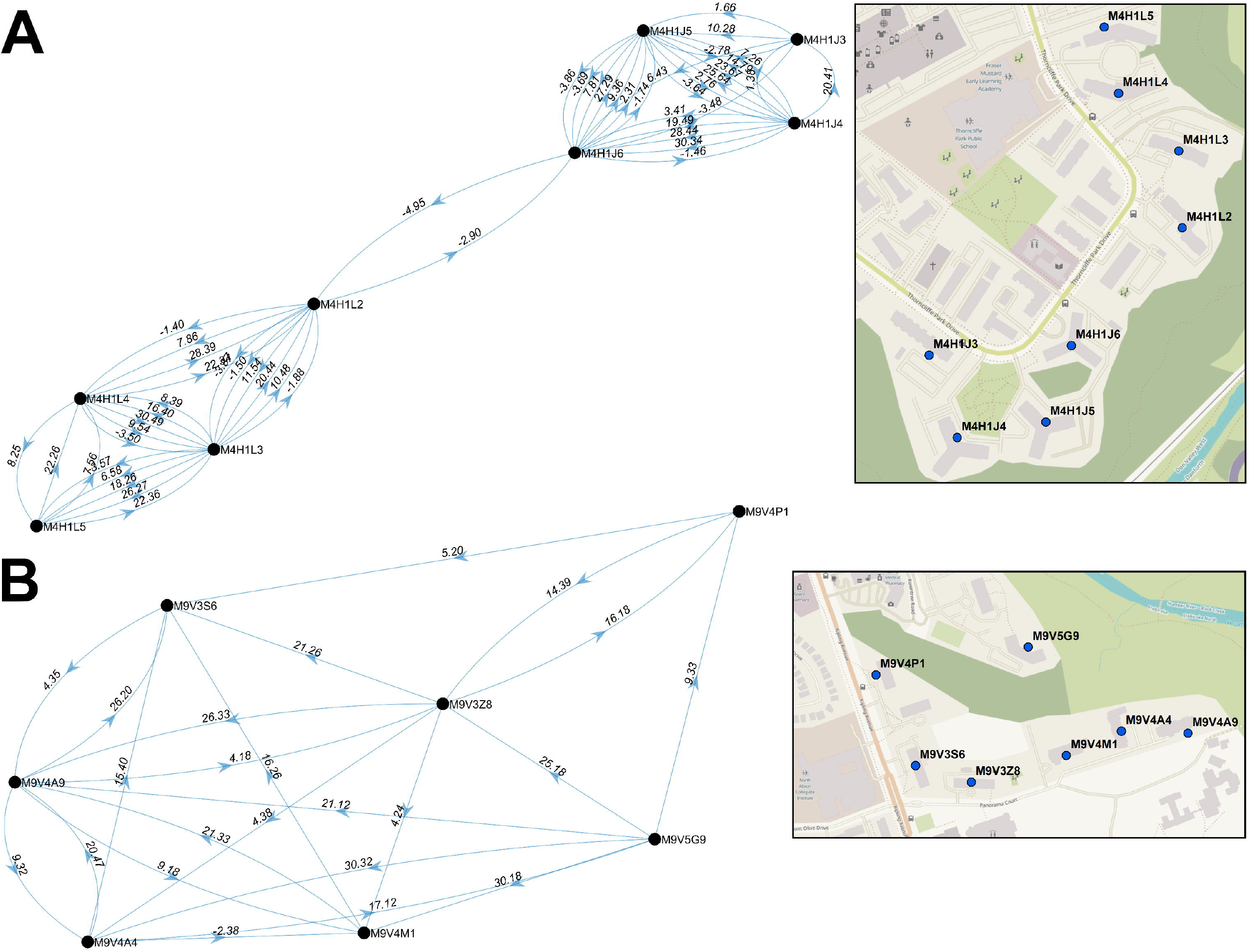
Multi-node Directional Networks of Closely Situated Clustered Postal Codes. Directional acyclic graphs (left) organize adjacent high-case clustered PC streaks within the same FSAs ordered according to their occurrence. Panels indicate PCs within (A) M4H and (B) M9V in Toronto during wave 2. Each connection (or ‘edge’) linking two PCs represents a pair of clustered streaks occurring within 30 days of each other (all significant PC pairs are indicated in Extended data^29^, Section 1 - Table S6). Labels on each edge indicate the duration (in days) separating adjacent PC pairs and the number of cases which occurred in the combined streaks (“dates.cases”). Negative values, when present, indicate the number of overlapping days of concurrent streaks in pairs of PCs. Arrows indicate the temporal order of these paired streaks (from earlier to the later streak). Maps (right) show the physical locations of each PC within the corresponding networks.

The multi-node networks during wave 3 consisted of only three postal codes (M3C, M9V, M9W; Extended data^29^, Section 1 - Figure S9A). Increasing the number of neighbors (by relaxing distance constraints, from 10 to 30 PCs) for these PC networks results in multi-node networks that were previously identified as bimodal (M4H, M4C, and M4X; Extended data^29^, Section 1 - Figure S9B). This procedure also resulted in four additional bimodal pairs of PCs in wave 2 (Extended data^29^, Section 1 - Figure S8D); however, the total number of PCs comprising the multi-node networks in wave 2 was unchanged (Extended data^29^, Section 1 - Figures S8A-C).

## Discussion

Geostatistical analysis revealed the locations and relationships between COVID-19 hotspots, utilizing daily postal code-level case data in Ontario Canada from March 26^th^, 2020 through June 25^th^, 2021 (including Gi*, Spatial Autocorrelation, Clustering and Outlier analyses and interpolation by kriging). Previous efforts to integrate geostatistical and epidemiological data have not investigated COVID-19 at this level of geographic and temporal resolution. This approach more precisely localized COVID-19 hotspots, and enabled early detection of disease clusters, which could be exploited to prevent or limit further spread at or close to these hotspots. Concurrent or consecutive neighboring hotspots classified as persistent high-case clusters were flagged as possible evidence of disease transmission.

The COVID-19 epidemic in Ontario has been classified into three distinct waves of infections^34^. The recurrence of successive waves shortly after easing of public health measures strongly motivated development and implementation of novel strategies that anticipate the locations and timing of future hotspots. Network analysis demonstrated that these locations may form nuclei from which subsequent local outbreaks arise. COVID-19 clusters were considerably diminished in the older at-risk population during the third wave 3, most likely, because of province-wide vaccination compliance, which resulted in lower mortality in long-term care residences during this period.

The techniques implemented in this study could prove useful to assist with contact tracing of infectious diseases, which may be challenging to carry out in a timely fashion once community spread has already occurred. Contact tracing has been vital to identify potential sources of viral transmission from infected cases. Successful contact tracing is impacted by incomplete collection of information, specifically interviewees who provide imprecise information, omit critical interactions, or who are uncooperative^35^. The directional acyclic network graphs presented are consistent with transmission between individuals residing in multiple PCs in close proximity to each other. These tools could assist tracing investigations by focusing resources on the most likely locations where interactions between infected and uninfected persons occur. It may be possible to exploit observations of COVID-19 infections at consecutive, neighboring PCs to anticipate the occurrence and locations of subsequent infections. This was exemplified by the progression of cases along PCs on Bathurst Street, Toronto in wave 2 (Figure 8C). We suggest that these and similar patterns may be useful as features along with cellular phone mobility data from dates preceding the appearance of future hotspots for predictive machine learning models. Accuracy in predicting the timing of future hotspots, however, would be influenced by the lag between data of infection and presentation of symptoms and by compliance of symptomatic individuals in seeking testing. Application of geostatistical analyses and other technologies^36^ may overcome the challenges posed by non-compliance or erroneous COVID-19 information from infected persons.

We also identified persistent, clustered postal codes that recurred between waves. Neighboring PCs comprising high-case clusters over a series of consecutive or overlapping dates were proposed as evidence of COVID-19 transmission. Nine PC pairs across 4 separate regions in Toronto were flagged in both waves 2 and 3 suggested recurrence. Without viral sequencing data, it is not possible to unequivocally establish whether these local outbreaks were independent events or due to re-emergence of the same infection from a low-level reservoir of asymptomatic or persistent mild infections during interwave 2-3. The same type of analysis did not yield evidence of inter-wave COVID-19 transmission in the other municipalities evaluated. Because of their comparatively lower population densities and smaller geographic size, it is less likely that the case thresholds required to define a COVID-19 hotspot within a 3-day window can be met in other Ontario municipalities. Increasing the aggregation of cases from 3-to 7-day sliding windows might improve detection of likely COVID-19 transmission in these areas.

Unlike traditional geostatistical epidemiology, we did not determine the likelihoods of hotspots based on simulations of random distributions of the same number of cases in the analyzed region^14,37^, since background levels of COVID-19 were undetectable prior to the pandemic. Therefore, performing geostatistical tests on positive COVID-19 cases is valid statistically. FSA-level analysis on population-normalized case counts (cases per 100,000; where FSAs with <100 individuals were not analyzed) was also performed (March to November 2020)^29^. Analyses could not be normalized by population densities (cases per 100,000), since these data are not publicly available for individual PCs. However, the smallest geographic units defined by Statistics Canada, termed Dissemination Areas, encompass an average of 15 postal codes (consisting of 400-700 persons)^38,39^.

A seroprevalence study of COVID-19 antibodies in Canada has suggested that infection rates were up to 3 fold higher than detected by RT-PCR testing during the first wave^33^. Similar studies have also indicated underreporting in the U.S.^40^ and Europe^41,42^. This included asymptomatic and untested symptomatic infected individuals^41^ or who received negative results^43^. Our study assumes that the geographic and temporal distribution of unrecognized infections trends similarly with known cases. However, nonuniformity in the distribution of asymptomatic and undetected cases across Ontario could have altered the detection or localization of hotspots. It seems likely that significant hotspots detected by geostatistical methods that also fulfill minimum case count thresholds would seem to be less likely to present as false positives or be incorrectly localized.

Some geostatistical functions (e.g., Gi* and Cluster and Outlier analysis) could be computed rapidly (within seconds for a single day analysis), while others (e.g., intersection of kriging contours PC boundaries) required up to 10 minutes per date analyzed on the ICES research computing system. In order to scale these computations over the duration of this project, it was critical to automate analyses for Provincial and multiple Municipal jurisdictions for timely generation and interpretation of these results. In April 2021, the sponsor of our AHRQ project requested an analysis of recent COVID-19 count and location data (January-March 2021). We automated kriging of localized hotspots in Toronto, Windsor/Essex, Ottawa and London within 5 days from the receipt of the request. This entailed performing the software analysis, imaging the results as maps, and interpreting these results. The rapid turnaround of geostatistical analysis of ongoing infectious diseases could have practical value for time-sensitive, critical public health management decisions. These programs have been made as a publicly available package, reformatted as a user-friendly toolbox in ArcMap (the *Geostatistical Epidemiology Toolbox*)^30^.

FSA-level geospatial tests, such as Gi*, did not reveal the distributions of COVID-19 infections, a limitation of this approach. We attempted to mitigate this by integrating the results of multiple geostatistical methods at different levels of resolution (e.g., Spatial Autocorrelation determined if cases were clustered at the PC-level). The locations of individuals in the same FSAs and PCs were aggregated at the centroids of their respective boundaries. However, boundary shapes and areas covered by different FSAs or PCs can vary, even though the total populations within each are more similar. Rural FSAs can be larger than urban counterparts and are occasionally non-contiguous (e.g., the FSA N0P). While close proximity of COVID-19 positive individuals in high-density municipalities is more consistent with transmission, it was nevertheless feasible to detect hotspots in rural regions. However, high concentrations of cases are required to find statistically significant hotspots in these regions. Since we use PC centroids to assign cases, this also affects the contours defined by kriging analysis of rural regions. These are limitations of the imposed boundaries of FSAs and PCs and not of the geostatistical tests used. Due to privacy considerations imposed by data providers, we could not perform kriging analysis on exact locations of COVID-infected individuals. Kriging interpolation can overlap multiple FSAs (Extended data^29^, Section 1 - Figure S10), which can be advantageous over area-to-area geostatistical tests, as hotspots in close proximity that cross an FSA boundary could be missed with these tests.

However, FSAs with few or no cases that are adjacent to a local hotspot could mistakenly be implicated. We account for this by ignoring FSAs without case counts, however this effect can still impact interpolation accuracy. For example, the interpolated case count for the FSA N8H in late June 2020 was much higher than the ground truth (Extended data^29^, Section 1 - Figure S4B). Over the same period, 109 cases were reported over a 3-day period within N0P 2J0, which is immediately north of N8H. The high-case kriging contour detecting this event intersects the boundary of N8H, thus causing this discrepancy (Extended data^29^, Section 1 - Figure S4C).

In late 2020, Ontario announced prioritization of COVID-19 vaccinations beginning with those with the highest risk of contracting severe illness. The subsequent group included individuals residing in hotspot communities^44^. These hotspots were identified in FSAs with the highest counts per 100,000 population. However, cases are not always uniformly distributed within FSAs with high disease burden (Figure 4). This study illustrates the importance of identifying hotspots at a higher resolution and describes how this level of precision provides targeted information about transmission that is often not feasible with FSA-level analyses. To minimize high rates of community transmission in the future, we suggest that targeted vaccination campaigns focus on highly constrained, localized hotspots with high case counts at the earliest possible juncture (rather than broadly across a region or FSA). For example, a hotspot in London was detected by kriging over several weeks at levels below our pre-selected interpolated case thresholds (Figure 6). The results of this analysis surpassed these thresholds approximately one week before it was publicly revealed. Thus, targeting can achieve the necessary precision and at an earlier stage with these methods. Smaller targeted locations may also facilitate communication, promote compliance and implementation of individual public health measures.

## Data Availability

A data repository Zenodo Archive has been created for this study. However, this archive does not provide complete COVID-19 case counts. To ensure patient privacy, ICES stipulated that the counts in all regions (FSAs or postal codes) exhibiting between 1 and 5 daily COVID-19 cases be masked. Therefore, all case counts between 1-5 will appear as NA within the files of this archive. To obtain access to the complete dataset, you will be required to apply to ICES directly (https://www.ices.on.ca/).

http://doi.org/10.5281/zenodo.5585811

## Acknowledgements

Data acquisition and computing facilities used in this study was supported by ICES, which is funded by an annual grant from the Ontario Ministry of Health (MOH) and the Ministry of Long-Term Care (MLTC). Data and information compiled were also provided by MOH. This study was also supported by the Ontario Health Data Platform (OHDP), a Province of Ontario initiative to support Ontario’s ongoing response to COVID-19 and its related impacts. The work described was funded and performed under a contribution agreement between the Innovation for Defence Excellence and Security (IDEaS) program at the Department of National Defence and CytoGnomix. The analyses, conclusions, opinions and statements expressed herein are solely those of the authors and do not reflect those of the funding or data sources. No endorsement by ICES, OHDP, its partners, or the Province of Ontario is intended or should be inferred. The authors acknowledge ICES personnel: Refik Saskin, Minnie Ho, Diana An, and Ryan Godinho, Kamil Malikov and his staff at MOH, and Regine Lecocq at the Department of National Defence IDEaS program for support and facilitation of this project.

## Competing interests

PKR cofounded and BCS and EJM are employees of CytoGnomix Inc.

## Data Availability

A data repository “Zenodo Archive for ‘Likely community transmission of COVID-19 infections between neighboring, persistent hotspots in Ontario, Canada’” has been created (http://doi.org/10.5281/zenodo.5585811^29^). This archive contains the following underlying and extended data, organized across 4 sections. Section 1 and 2 contain extended data, and Sections 3 and 4 contains the data files containing and the map image files displaying the underlying statistics from the various geostatistical analyses performed in this study.

### Extended Data

Zenodo Archive for ‘Likely community transmission of COVID-19 infections between neighboring, persistent hotspots in Ontario, Canada’: http://doi.org/10.5281/zenodo.5585811^29^

This project contains the following data:

Section 1 – An Excel document containing 6 additional tables described in this study which provide summary statistics of various geostatistical tests (“Section 1 – Tables S1-S4”) and lists all identified single and paired high-case cluster streaks (“Section 1 – Tables S5-S6”). This section also contains 10 additional figures referred to in the manuscript (“Section 1 – Figures S1-S10”) with a Word document which describe them.

Section 2 – All localized hotspots (identified through kriging analysis) catalogued for each municipality evaluated are provided in this section. Each data file lists the FSA containing the localized hotspot, the size of this hotspot (% by area), the 3-day window in which it was observed, the interpolated and true case values on this date, and whether the FSA was also significant by Gi* analysis (on any of the 3 days analyzed).

### Underlying Data

Section 3 – All output files from the geostatistical tests performed in this study are provided in this section. This includes the output from Ontario-wide FSA-level Gi* and Cluster and Outlier analyses, and PC-level Cluster and Outlier, Spatial Autocorrelation, and kriging analysis of 6 municipal regions. Kriging analysis output files are also provided for 7 municipalities immediately surrounding Toronto. This section also contains data files from our analyses of stratified case data (by age, gender, and at-risk condition). All coordinates presented in these data files are given in “PCS_Lambert_Conformal_Conic” format. Case values between 1-5 were masked (appear as “NA”).

Section 4 – Sets of image files which map the results of our geostatistical analyses onto a map of Ontario or within the municipalities evaluated are provided. This includes: kriging analysis (PC-level), Local Moran’s I Cluster and Outlier analysis (FSA and PC-level), normal and space-time Gi* analysis, and all images for all analyses performed on stratified data (by age, gender and at-risk condition).

### Software

A second Zenodo repository (doi.org/10.5281/zenodo.5675978^30^) provides all program files pertaining to the *Geostatistical Epidemiology Toolbox* (Geostatistical analysis software package to be used in ArcGIS), as well as all other scripts described in this manuscript. This code is available under the terms of the GNU General Public License v3.0.

## Extended Data Section 1 – Supplementary Figure Legends

(legends are described in Zenodo archive doi.org/10.5281/zenodo.5585811^29^)

**Extended Data Section 1 - Figure S1. Case Count of FSAs by Frequency**

Histograms indicate the frequency in which an FSA in (A) Ontario and (B) Toronto were reported to have ≥X COVID-19 cases (where X is the value on the X-axis) on a specific day from March 1^st^ to September 24^th^, 2020 (blue columns). Gi* analysis identified which FSAs were significant hotspots by comparing them with neighboring FSAs at a fixed distance (indicated with red bars) and inversely weighted distances (indicated with green bars), at a threshold of <50km. A large fraction of FSAs in Ontario and Toronto contained fewer than 8 cases and were not statistically significant. Over two thirds of FSAs with at least 10 cases were significant by the Gi* test at a 99% confidence level.

**Extended Data Section 1 - Figure S2. Evaluation of Kriging Semivariogram Models**

Interpolation by different semivariogram models was evaluated using kriging to determine which model best approximated true case counts in regions of moderate to high population densities. The two best performing models chosen were ‘Power’ and ‘Thin Plate Spline’ (TPS). Each point in this figure represents PCs with ≥1 COVID-19 case and PCs with ≥6 cases have been labelled [PC / case count]). Panel (A): Kriging analysis of Hamilton (moderate-density of cases) indicates Power-modeled kriging (left) shows smooth fit to the actual case distribution compared to TPS, which is discontinuous (right). Panel (B): In Toronto (high-density of cases), interpolation with the ‘Power’ model more frequently under-estimated actual case counts, whereas the TPS model derived case counts that more precisely resembled the actual distribution of cases.

**Extended Data Section 1 - Figure S3. Histograms of the Fraction of FSAs Overlapped by Significant Kriging Contours**

The percent overlap of the areas covered by kriging-derived contours and the corresponding FSAs were computed by area for all FSAs flagged by Gi* analysis. Panels depict (A) wave 1, (B) interwave 1-2, (C) wave 2, (D) interwave 2-3, and (E) wave 3. Histograms illustrate the distribution of the fraction of each FSA that is geographically intersected with either the highest interpolated kriging contour level (left) or kriging contours interpolating at least 5 (right).

**Extended Data Section 1 - Figure S4. Hotspots Localized within High Disease Burden Regions**

The distribution of COVID-19 cases within the FSAs (A) N9Y (Kingsville, ON) and (B) N8H (Leamington, ON) during interwave 1-2 was evaluated by kriging, Spatial Autocorrelation and Gi* analysis. Kriging- interpolated case values (blue line) were similar to actual case counts (red line). COVID-19 outbreaks within these FSAs were often identified by Gi* analysis (green asterisks), however the distribution of cases within these regions were infrequently found to be spatially clustered (purple crosses). Panel (C) indicates Power-based kriging contours can be large in regions with low population densities due to the distribution of cases. Interpolation of cases within N8H from June 22^nd^ to June 26^th^, 2020 exceeded than actual counts due to the contribution of a high-case postal code in a neighboring FSA (N0P), with the resultant kriging contour intersecting both FSAs.

**Extended Data Section 1 - Figure S5. A Localized Hotspot in Leamington, Ontario**

PC-level COVID-19 case data of the FSA: N8H (Leamington, ON) on consecutive dates in December 2020 evaluated by kriging interpolation (blue line), testing for significance by Spatial Autocorrelation (purple crosses) and Gi* analysis (green asterisk). B) Map contours and distribution of cases. Small circles in this region represent PCs with one or more cases from December 21^st^ to the 23^rd^, 2020. The maximal kriging contour was coincident with the hotspot in PC N8H 3V5 during this window.

**Extended Data Section 1 - Figure S6. Spatial Clustering and the Identification of Localized Hotspots**

Spatial autocorrelation analysis identifies clusters of postal codes with high case counts within FSAs. Panels (A) displays significantly clustered cases in Toronto FSA M1P from Nov. 3-5, 2020. This region was also significant by Gi* and contained a localized hotspot consisting of 8 cases (M1P 4V6). Each dot represents a postal code with at least 1 but fewer than 6 cases. (B) Cases in M1C were also significantly clustered from Nov. 6-8, 2020, but no hotspots were identified. Topographic contours indicate kriging-interpolated case counts.

**Extended Data Section 1 - Figure S7. Contiguous or Consecutive Toronto Paired Streaks considering 50 Closest Neighbors**

High-case postal code clusters were identified by Cluster and Outlier analysis (Local Moran’s I). High-case cluster streaks (PCs significant over multiple consecutive days, allowing for a single day gap) were identified and paired to other streaks within close proximity (considering 50 closest neighbors based on distance between PC centroids). Each point represents a paired PC streak with colors representing the total number of cases in both, the X axis indicates the duration between PC pairs (with negative values representing degree of overlap between end of first streak and beginning of the other), and the Y axis indicates spatial distance between the centroids of each PC forming a pair. All paired streaks are illustrated, regardless of wave.

**Extended Data Section 1 - Figure S8. Directional Acyclic Graph Network of Clustered Streaks in Toronto during Wave 2**

Directional networks identify streaks of clustered high-case counts (≥ 6 cases for each date evaluated) in close spatial and temporal proximity. Each connection (or edge) represents two neighboring PCs, where the value preceding the decimal represents the days separating pairs of streaks followed by the total number of cases in both (“dates.cases”). The arrows represent the order in which streaks occur in the pair of PCs. The left and right panels indicate the same network, based on either the 10 or 30 closest neighbors, respectively. Maps indicate the locations of PCs within the network (middle). The panels highlight different networks in neighborhoods according to the FSA: in which they occur: (A) M4H, (B) M9V and (C) M2R. Although increasing the number of neighbors considered for pairing added new edges, novel postal codes were not introduced. (D) Networks only connecting streaks between a single pair PCs are indicated.

**Extended Data Section 1 - Figure S9. Directional Networks of COVID-19 Clustering Streaks in Toronto during Wave 3**

Directional networks illustrate the relationship between PCs with consecutive COVID-19 clustering (i.e., streaks) in Toronto during Wave 3 of the COVID-19 epidemic, considering the (A) 10 closest and (B) 30 closest neighboring PCs for each streak. Simple networks involving two or three PCs were common and exhibited recurrent transmission between nodes. Increasing the number of allowable neighbors considered increased network complexity, including those not previously seen during Wave 2 (M3C, M4C and M4X). Networks for Wave 3 are less complex compared to those in Wave 2, possibly due to the increase of vaccinated individuals within the population.

**Extended Data Section 1 - Figure S10. Overlap of Localized Hotspots with Multiple FSAs**

Kriging contours can overlap multiple FSAs. While the kriging semivariogram model selected can affect their frequency, contours which intersect ≥2 FSAs can occur with either method. Panel A includes: a TPS-based kriging contour which intersected 9 separate FSAs in Toronto, and panel B shows a power-based kriging contour which interpolated ≥10 cases over 6 unique FSAs in London.

## Notes

### Author Declarations

Access to anonymized Ontario COVID-19 test results through the ICES Research and Analytic Environment (RAE) was obtained through Applied Health Research Questions (AHRQ) project #P0950.096 approved by the Ontario Ministry of Health and Long-Term Care (MOHLTC). ICES has been designated to receive and use data under the province's Personal Health Information Protection Act. All authors have completed privacy training and signed a data access agreement with ICES.

